# Return-to-work for People Living with Long COVID: A Scoping Review of Interventions and Recommendations

**DOI:** 10.1101/2024.12.10.24318765

**Authors:** Gagan Nagra, Victor E. Ezeugwu, Geoff P. Bostick, Erin Branton, Liz Dennett, Kevin Drake, Quentin Durand-Moreau, Christine Guptill, Mark Hall, Chester Ho, Pam Hung, Aiza Khan, Grace Y. Lam, Behdin Nowrouzi-Kia, Douglas Gross

## Abstract

**Introduction:** Long COVID affects individuals’ labour market participation in many ways. While some cannot work at all, others may return to work (RTW) in a limited capacity. Determining what rehabilitation or related strategies are safe and effective for facilitating RTW is necessary.

**Objectives:** To synthesize evidence on RTW interventions for people living with Long COVID and to identify ‘promising’ interventions for enhancing work ability and RTW.

**Methods:** We followed Arksey & O’Malley’s methodology and the PRISMA extension for scoping reviews. Five electronic bibliographic databases and grey literature were searched. The included various study designs, such as randomized controlled trials (RCT), quasi-experimental designs, and observational studies. Two reviewers conducted screening and data extraction, with disagreements resolved through consensus. Intervention studies were categorized as promising (statistically significant RTW outcomes or ≥ 50% RTW), somewhat promising (20% to < 50% RTW), or not promising (non-statistically significant RTW outcomes or < 20% RTW).

**Results:** Eleven recommendations and eleven intervention studies were identified. Of the intervention studies, 6 were cohort studies, 3 quasi-experimental studies, 1 RCT and 1 case report. Promising interventions included multimodal and interdisciplinary work-focused rehabilitation (1 article), psychoeducation, pacing, and breathing strategies (2 articles), shifting focus from symptom monitoring to optimizing functional outcomes (1 article), and enhanced external CounterPulsation (EECP) inflatable pressure to improve blood flow (1 article).

**Conclusion:** Many uncertainties remain regarding which RTW interventions are effective or the optimal characteristics of these interventions.

## 1. BACKGROUND

Post-COVID-19 condition (PCC), or Long COVID, is characterized by the presence of new onset or persistent symptoms 3 months after a suspected or confirmed history of SARS-CoV-2 infection (1–3). Long COVID is a complex and multi-faceted condition that affects individuals in different ways (4). About 16% of Canadians infected with COVID-19 develop Long COVID, with higher prevalence in women, individuals hospitalized due to COVID, those with pre-existing health conditions or repeated COVID-19 infections (3). Long COVID symptoms limit daily functioning, work ability, and employment, with up to 76% of people with Long COVID taking time off work or stopping work entirely (3, 5–7). Previous qualitative research highlighted significant challenges faced by individuals with Long COVID in performing daily activities and meaningful work-related tasks (8, 9). About 3.5 million Canadian adults reported experiencing long-term symptoms following a COVID-19 infection with about 100,000 reporting they were still off work due to symptoms as of June 2024 (3, 10), which corresponds to approximately 0.45% of the Canadian labour force.

Systemic exertion intolerance, characterized by severe fatigue worsened by physical or mental exertion, is a prevalent symptom in Long COVID and can severely impact sustained work activity (1, 5, 11). Factors like exercise, stress, cognitive tasks, and sleep deprivation can exacerbate symptoms of systemic exertion intolerance (3, 5). This prevalent symptom is similar to Myalgic Encephalomyelitis (ME), and some individuals with Long COVID meet diagnostic criteria for ME (1, 11–13). Other challenges faced by people with Long COVID include fluctuating and episodic symptoms, sleep disruption, and fatigue, hindering daily activities and return to work (RTW) (3). Traditional therapies like graded exercise may not be effective or could be harmful for some people with Long COVID, particularly in those experiencing Post-Exertional Malaise (PEM), necessitating exploration of new strategies (14). Despite recognition of the burden of Long COVID, research on strategies to improve daily function and work ability is limited.

Rehabilitation is one option for potentially addressing Long COVID activity limitations and participation restrictions due to its role in decreasing the burden of physical, neurocognitive, and psychological limitations with the aim to stabilize or enhance patients’ physical and mental abilities (15). However, there are few Long COVID-specific rehabilitation tools or protocols to address various symptoms such as fatigue or cognitive dysfunction (16). This can lead to diverse and inconsistent intervention approaches or even potentially harmful solutions such as graded exercise, which is now contraindicated for those experiencing PEM (3, 17). Research is still inconclusive around other exercise-based rehabilitation interventions (7, 17).

Determining what rehabilitation strategies are safe and effective for facilitating RTW is necessary. There is an urgent need for safe, effective and evidence-based Long COVID treatment programs (16). Some studies of rehabilitation strategies have begun to emerge, although the literature is diverse, emerging and requires synthesis (8, 15, 18, 19). This could help to inform rehabilitation practice guidelines globally. The purpose of this scoping review is to synthesize existing evidence about Long COVID interventions and their impact on RTW. We sought to identify promising strategies to promote work activity (return to or stay at work) and explore impactful RTW recommendations for individuals living with Long COVID.

## 2. METHODS

### Study design

Searching and reviewing the literature revealed that a scoping review had not previously been conducted solely on the impact of rehabilitation strategies and programs on the outcome of RTW for people with Long COVID. For this scoping review, the Arksey & O’Malley methodological framework was applied, and reported based on the PRISMA extension for scoping studies (20). This methodological framework consists of the following main stages: identifying the research question, identifying relevant studies, study selection, charting the data, collating, summarizing, and reporting results, and consultation (optional stage) (20). This methodology guided this scoping review to summarize and disseminate research findings to determine what is known about Long COVID interventions and their impact on RTW.

We followed the PI(E)COS framework (population, intervention, exposure, comparators, outcomes, study design/setting) and included various study designs, such as randomized controlled trials, quasi-experimental designs, and observational studies.

### Search terms and search strategies

Members of the research team met with a health sciences librarian (LD) to do preliminary searches and develop an extensive list of relevant terms. The librarian then conducted a systematic search of MEDLINE (via Ovid), Embase (via Ovid), APA PsycINFO (via Ovid), CINAHL Plus with Full Text (via EBSCOhost), and Cochrane Library CENTRAL trials (via Wiley) from database inception until October 6, 2023. The search strategy included a combination of subject headings and keywords to combine the concepts of Long COVID and either occupational rehabilitation or occupational functioning/outcomes. The search was optimized for running in each database. No date, language or publication type limits were applied to the search.

In addition to searching academic databases for peer reviewed literature, an extensive grey literature search was conducted. The librarian searched on Nov 7, 2023 the following websites and online databases: MedRxiv (https://www.medrxiv.org/), Bielefeld Academic Search Engine (https://www.base-search.net/), OAIster (oaister.on.worldcat.org), Custom Google Search Engine for Canadian Public Health Information (https://www.ophla.ca/p/customsearchcanada.html), Custom Google Search Engine for US State Government Information (https://www.ophla.ca/p/customsearchusstates.html), Government of Canada Publications (https://www.publications.gc.ca/site/eng/home.html), Alberta Health Services Long COVID page (https://www.albertahealthservices.ca/topics/Page17540.aspx), WHO pages relevant to Long COVID and a number of general searches on Google. The full search strategy is available in Appendix A.

#### Selecting studies for analysis

The following were the final set of inclusion/exclusion criteria for the review:

*Topic of the article* – Evaluation of an intervention aimed at increasing work functioning or work ability.

*Population* – People living with Long COVID. Our review included people with a variety of symptoms and activity limitations due to Long COVID. Given the diverse nomenclature in this area, a variety of search terms were included.

*Intervention* – We focused primarily on studies evaluating interventions aimed at improving work functioning or facilitating RTW. Functional recovery is a crucial outcome in Long COVID. From the perspective of the various stakeholders involved (i.e. people living with Long COVID, workers’ compensation insurers, employers and health care providers), work-related functional recovery — such that the patient can return to sustainable and predictable work activity — is important and has important career and quality-of-life implications.

*Outcome* – Work-related outcomes, including RTW, stay at work, work disability, work absence/absenteeism, and work productivity or related constructs (i.e., presenteeism, etc).

*Study type* – Any design evaluating an intervention for Long COVID. Systematic reviews were excluded but references within those located were searched for further articles.

#### Screening of relevant articles

The titles and abstracts of articles obtained from the online databases were reviewed and appraised for relevance. Two researchers from the team read each title/abstract independently and judged whether they were relevant to the research question. When there were disagreements between reviewers, the principal researcher (DPG) offered additional consultation until a consensus could be reached. If the relevance of a study was still unclear, then the full article was obtained. After identifying relevant abstracts and titles, two independent researchers assessed the corresponding full versions of the studies to determine which articles were relevant for inclusion in the full review. All team members used Covidence software (Melbourne, Australia) to organize data at all stages of this review.

#### Consultation with People With Lived Experience and Knowledge Users

The consultation process for this study included a large multidisciplinary team consisting of people with lived experience of Long COVID impacting work ability, researchers from various health professions (occupational medicine, physiotherapy, occupational therapy, and kinesiology), and knowledge users who were experienced clinicians providing rehabilitation for Long COVID or employees of the Workers’ Compensation Board of Alberta. Meetings were held with the full team, then periodic email updates were provided to seek advice on selecting studies for analysis and to summarize and report results. Knowledge users were asked whether they knew of any interventions that were currently in use and found to be safe and effective in the Long COVID population. Feedback from knowledge users highlighted the importance of: 1) considering the variable and episodic nature of Long COVID symptoms as a major barrier to RTW, 2) including RTW outcomes as search terms; 3) considering not only papers describing specific interventions, but also theoretical or conceptual papers dealing with models or pathways for people living with Long COVID; and lastly, 4) considering the importance of safety, feasibility, burden, and need for training in addition to scientific validation when considering the interventions identified from the literature search. Before charting the data, the knowledge users were consulted to determine whether the number of articles selected was appropriate and whether the search terms should be altered.

### Data Analysis

#### Charting the Data

Reviewers extracted relevant information from each article and entered it into Covidence. This included the author list, year of publication, article title, geographic location of the study, type and brief description of the intervention, study population, study design and goals, methods used, outcome measures used, important results and any economic data recorded.

#### Collating, summarizing and reporting results

During this stage, we created an overview of all research located. Initially, we presented a basic numerical summary of the studies, including the extent, nature and distribution of the articles. Then, we summarized articles according to the types of tools described or evaluated, research methods used, populations studied, and study results/outcomes. Since the scoping review methodology was intended to summarize both the breadth and depth of the literature, we reported the number of articles for various interventions as well as descriptive information about the articles.

We attempted to map the diversity of studies observed to create an inventory of the various study designs and methods used. This procedure allowed us to draw conclusions about the nature of research in this area and provide recommendations for future studies. The various interventions identified in the articles were categorized, and key concepts and terminology used in the articles were summarized in the Appendix (see Appendix A). A descriptive characteristic summary and content analysis of intervention studies were conducted to collate, summarize, and report results for this scoping review (20).

## 3. RESULTS

Reliability of the screening process of screening titles, abstracts, and full texts was high with an average agreement percentage of 81% between reviewers. The initial search of the online databases identified 2,056 potentially relevant articles. Once duplicates were removed, 1,374 unique studies were included for the screening of titles, abstracts, and full texts. The full texts of 177 articles were screened and from that process 11 research articles and 11 relevant guidelines were found. Despite an extensive internet-based grey literature search and reviewing the reference lists of systematic reviews, no additional studies were located by these methods. Thus, 22 relevant articles were included for data extraction. Figure 1 shows the PRISMA flow chart of our article search and relevance selection process. A search of the grey literature obtained no new documents or websites describing evaluations of interventions promoting RTW in people living with Long COVID.

**Figure 1:**
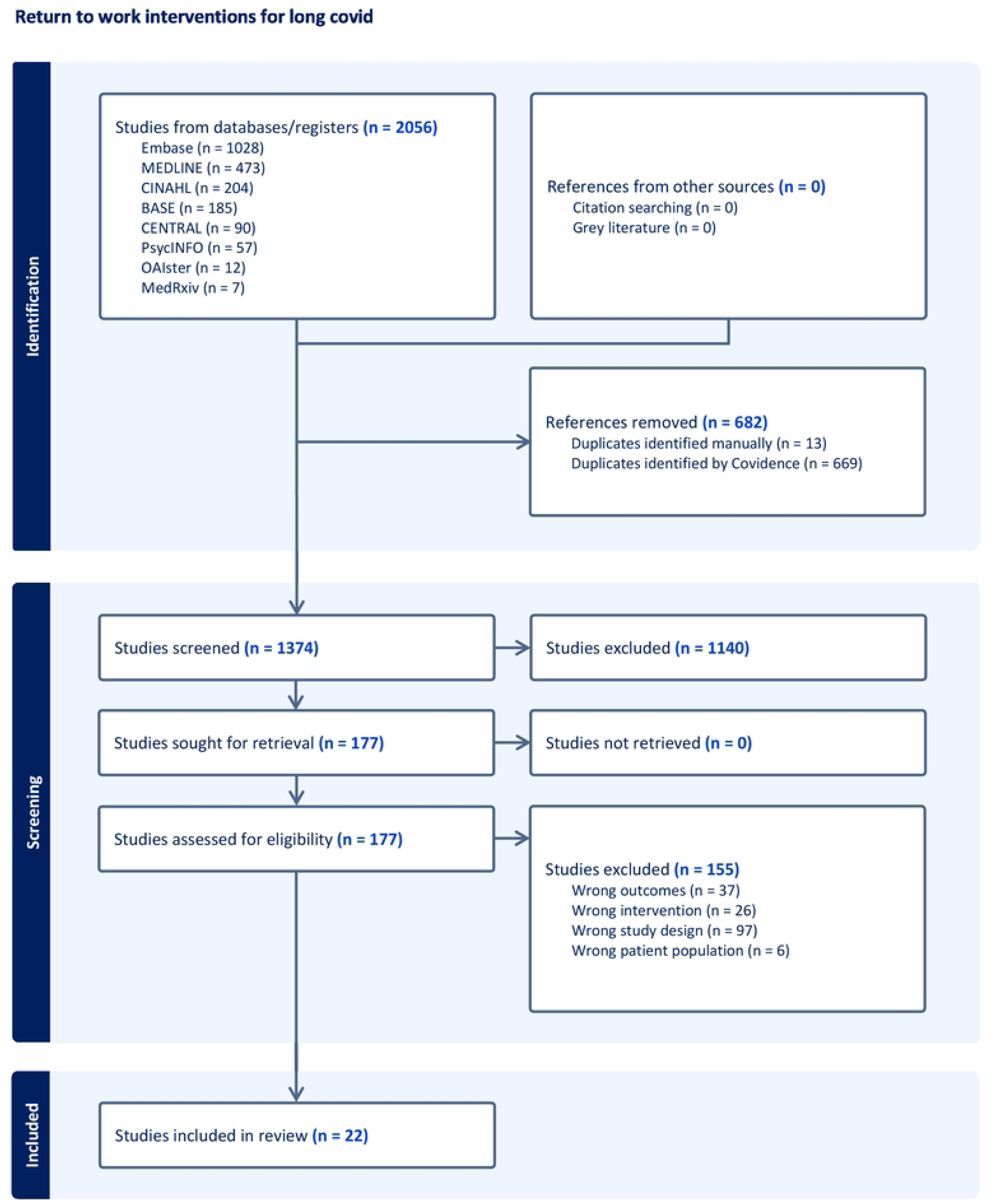
PRISMA flow diagram

### 3.1. Descriptive summaries of included studies

Eleven relevant intervention studies were included (see Table 1, Appendix B). Furthermore, a descriptive summary of the intervention studies including intervention details, results, and proportion of RTW was conducted (see Table 2, Appendix C). A summary of key messages for RTW recommendations from eleven clinical practice guidelines (see Table 3, Appendix D) was also created to synthesize findings identified in the literature (20). Summaries were organized based on relevance to the study question.

**Table 1.**
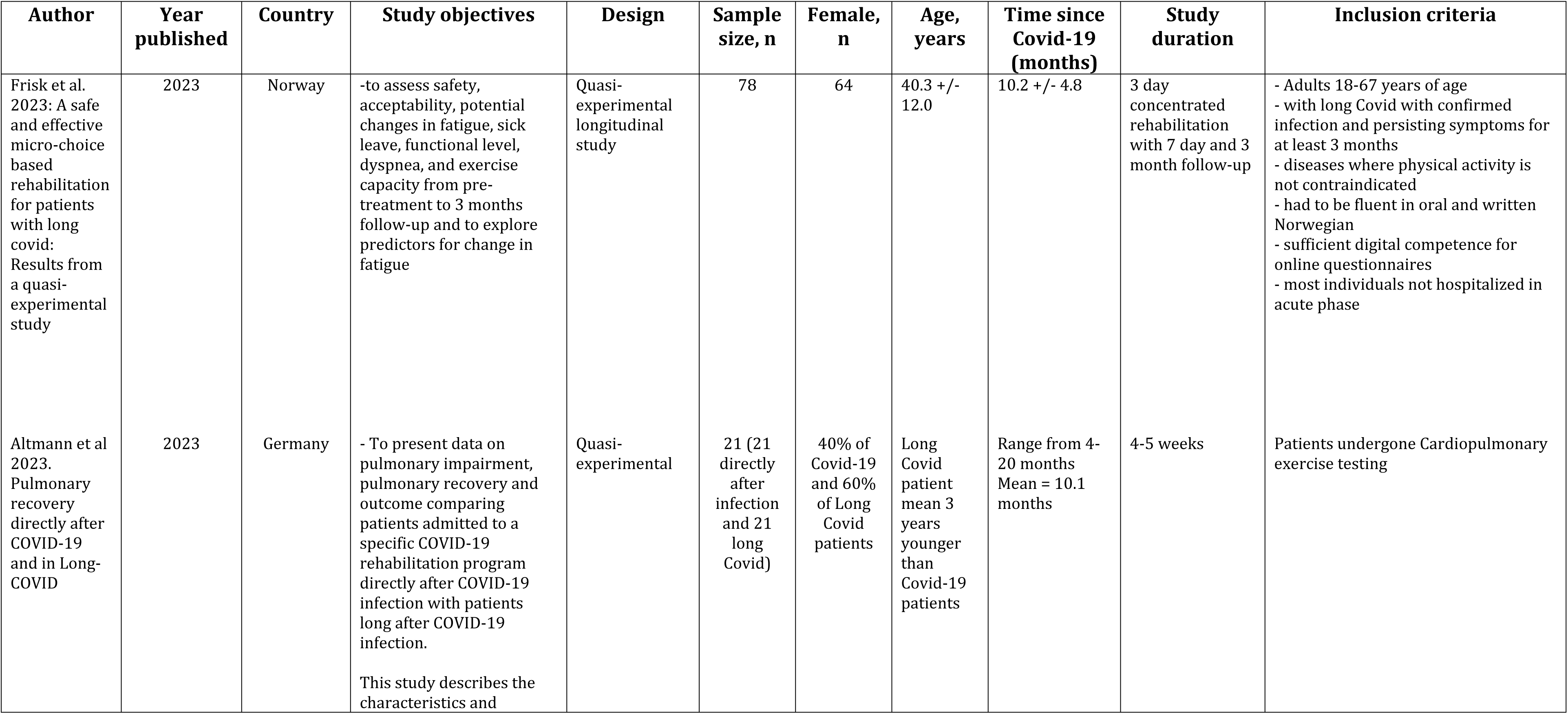
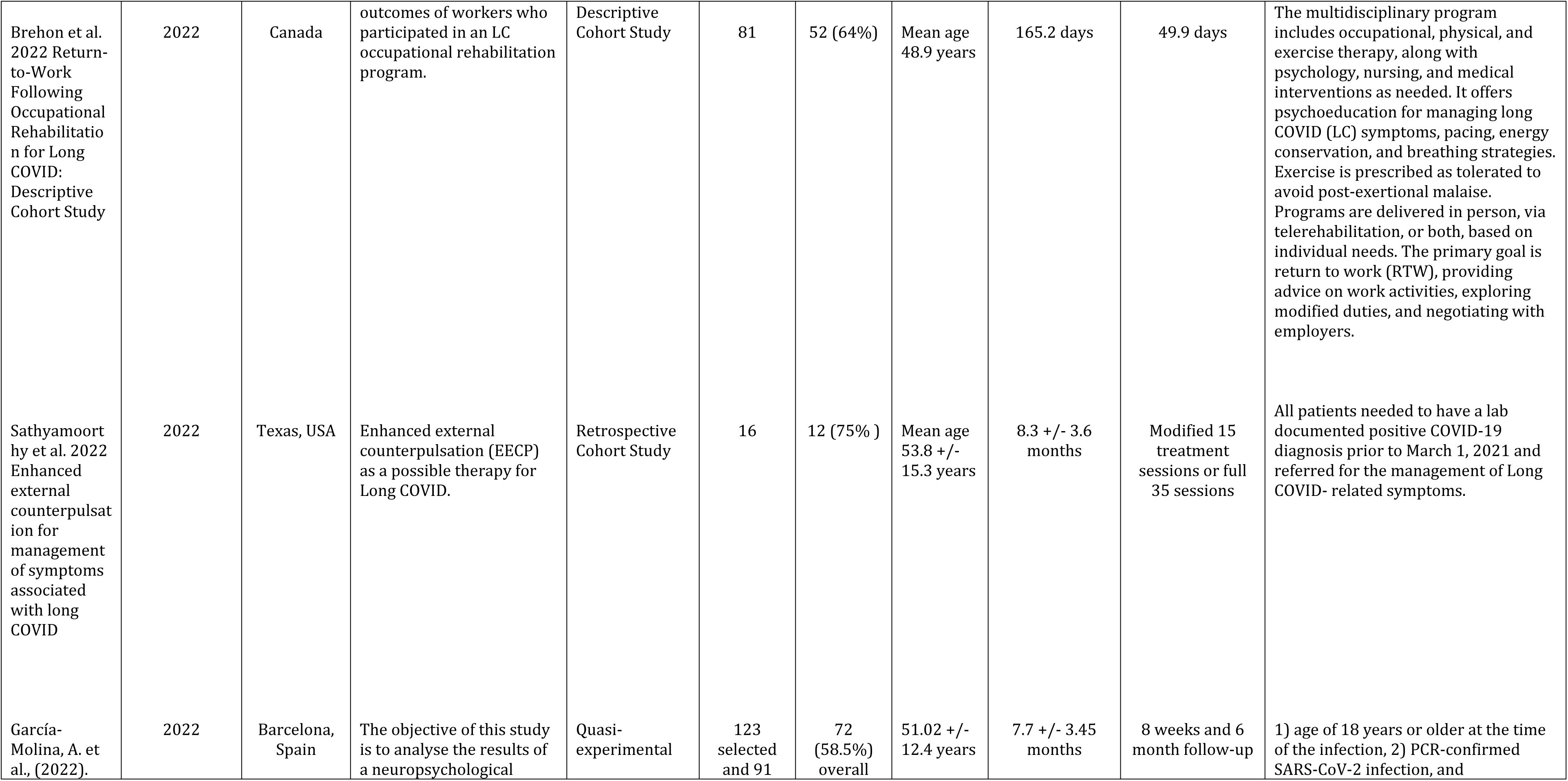
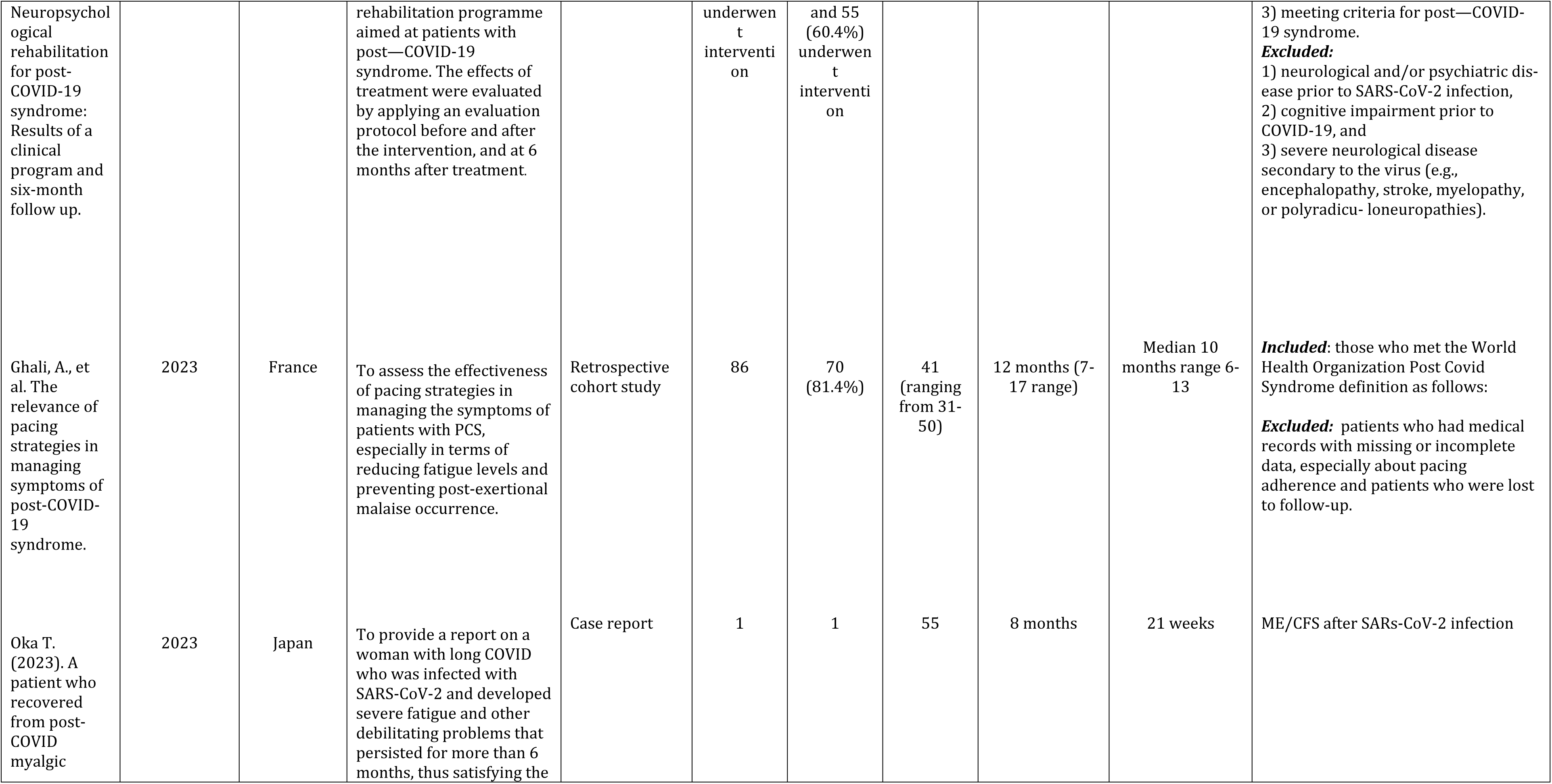
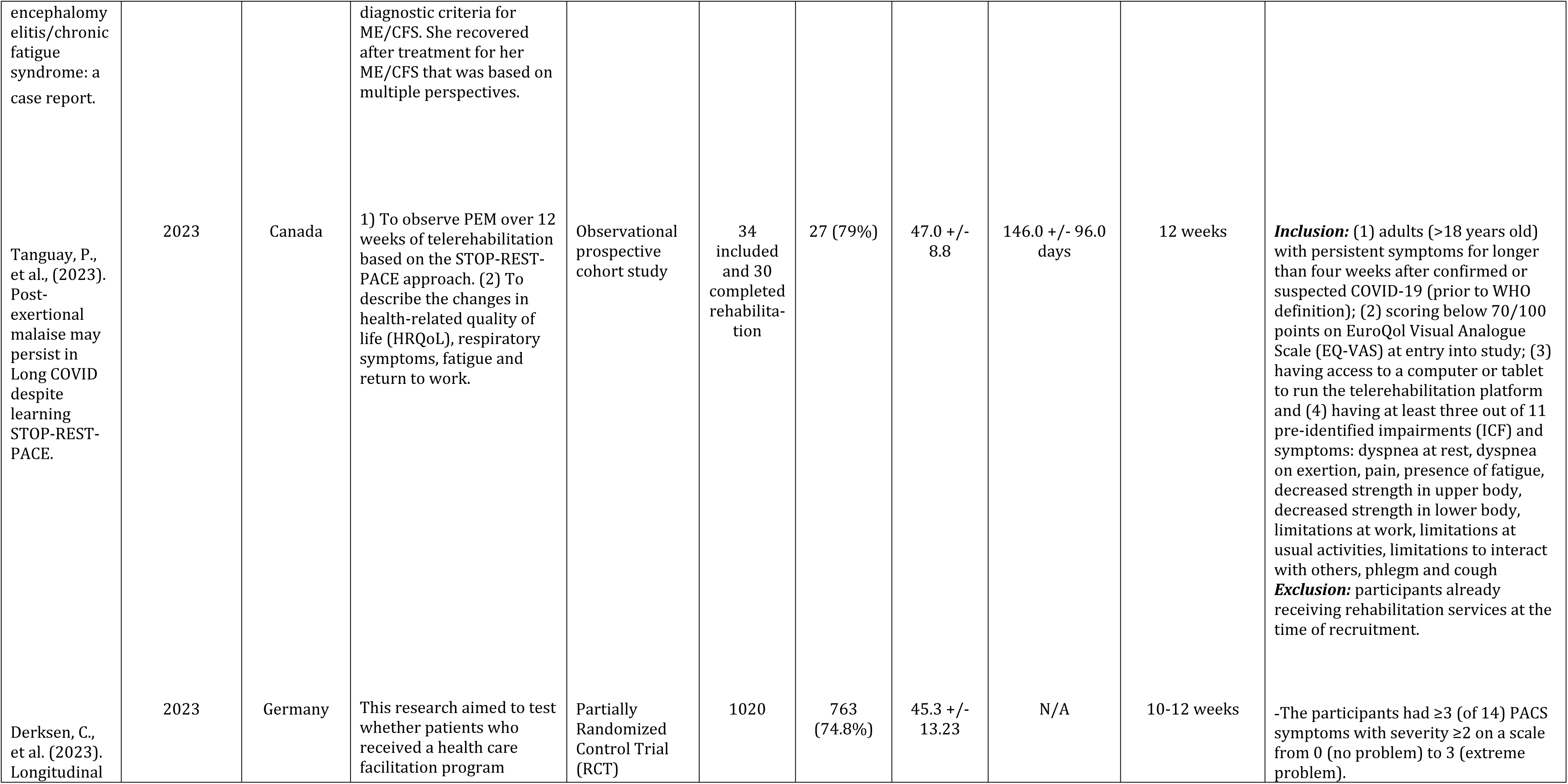
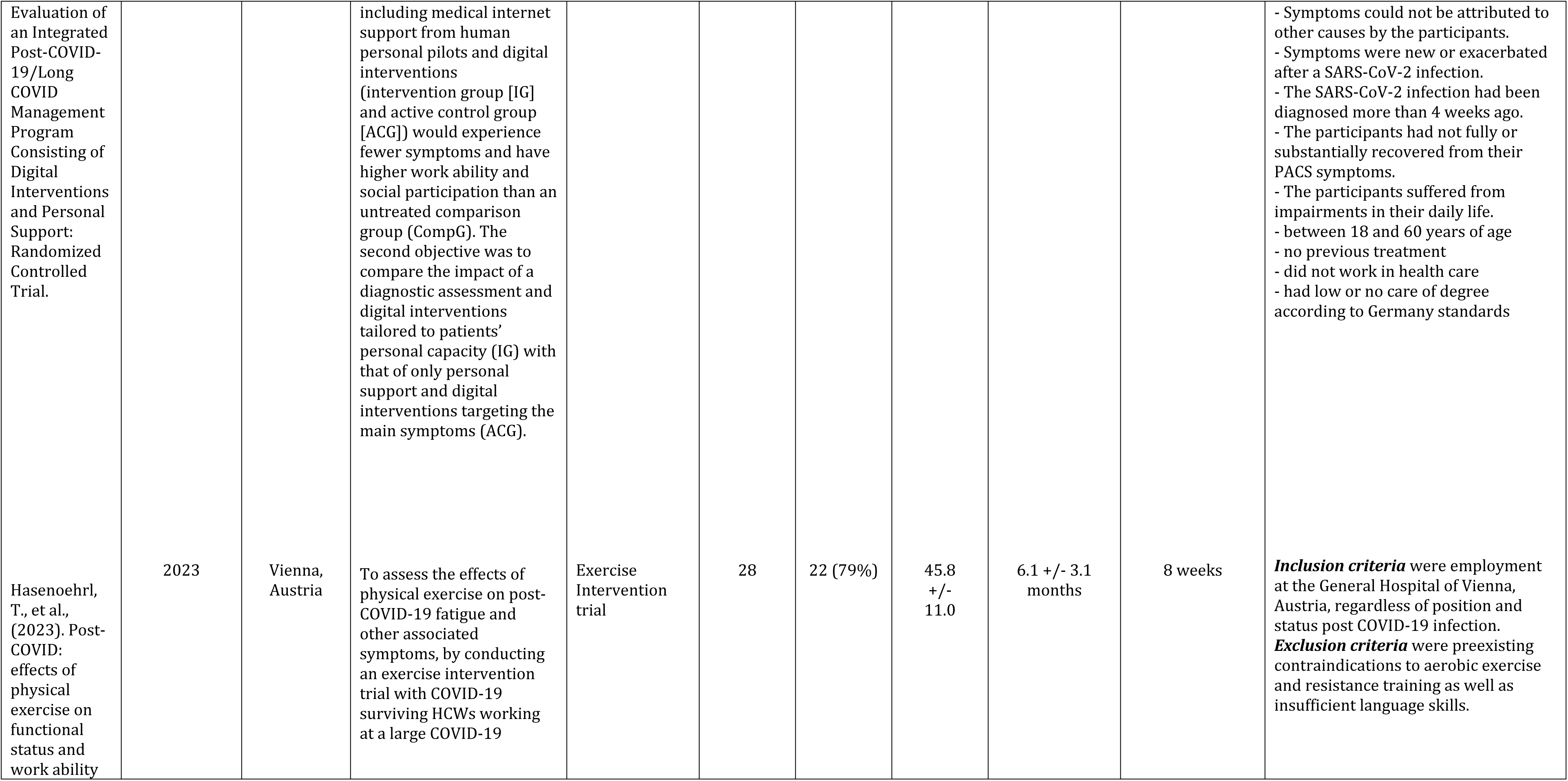
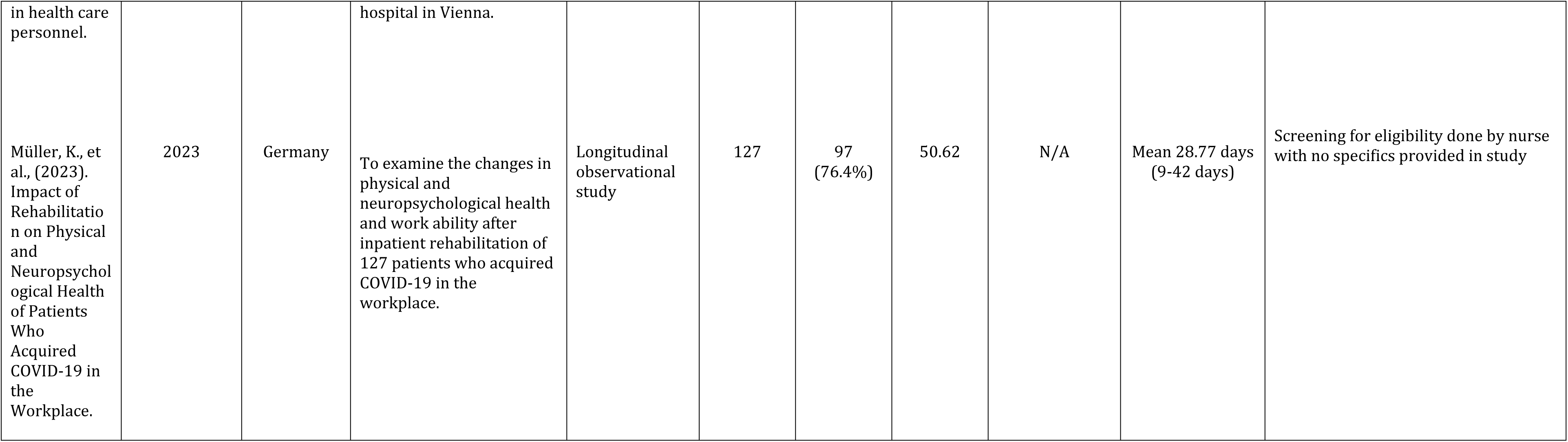
Descriptive Characteristics of Intervention studies.

**Table 2:**
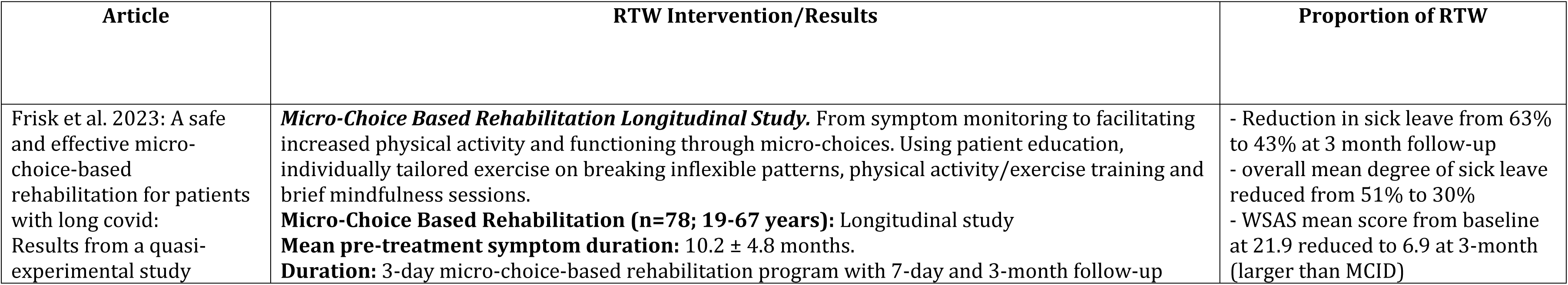
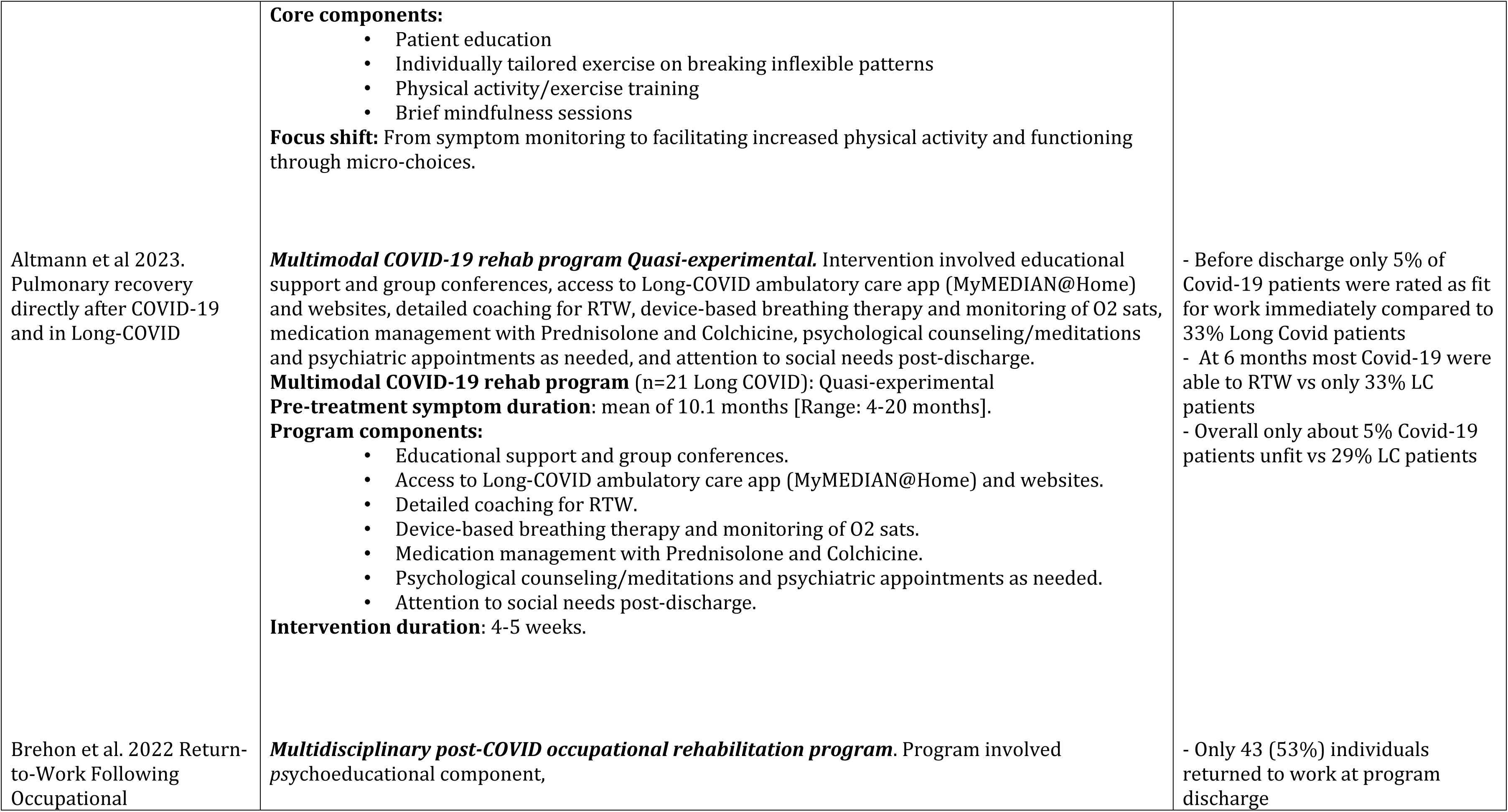
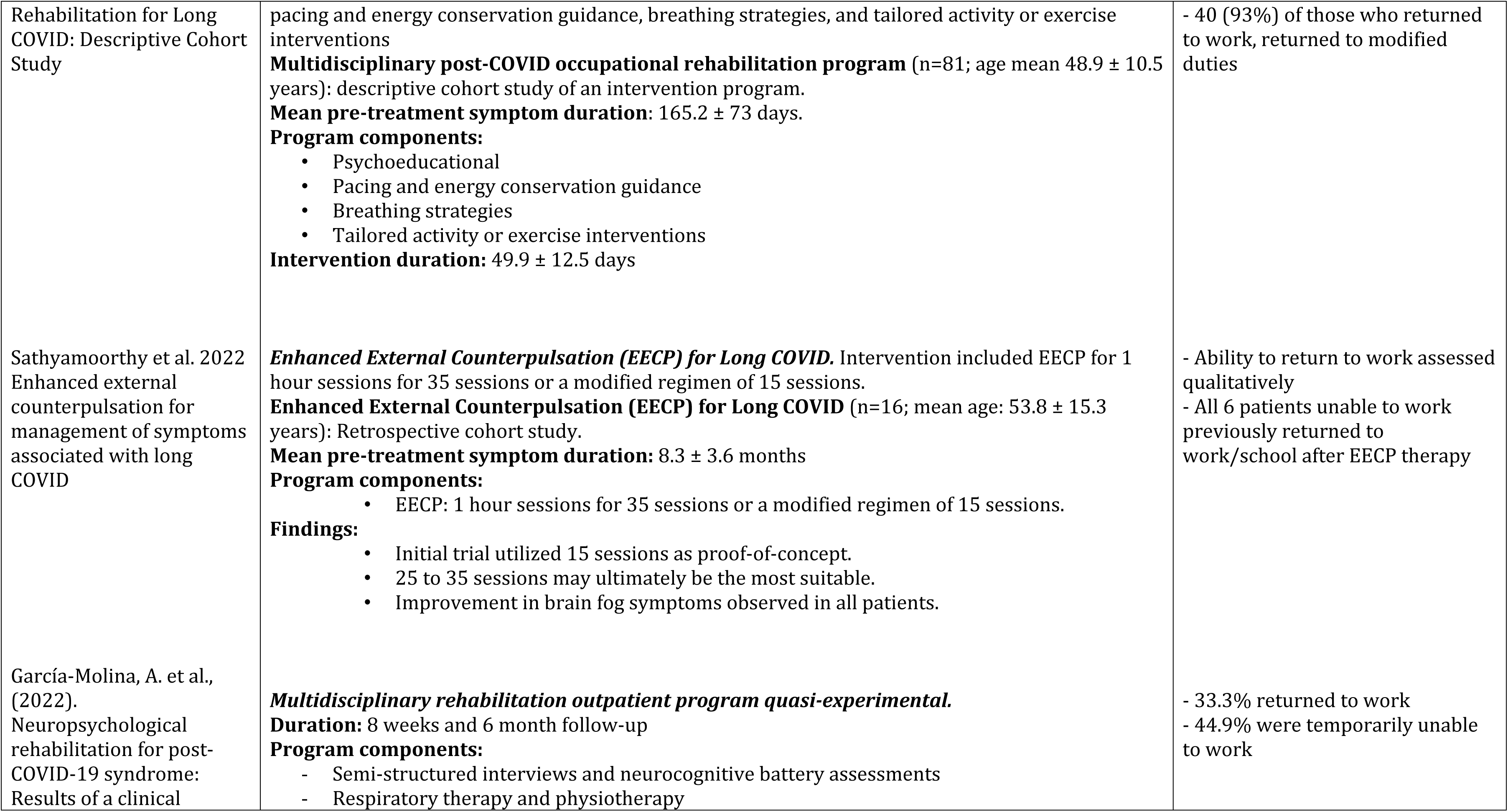
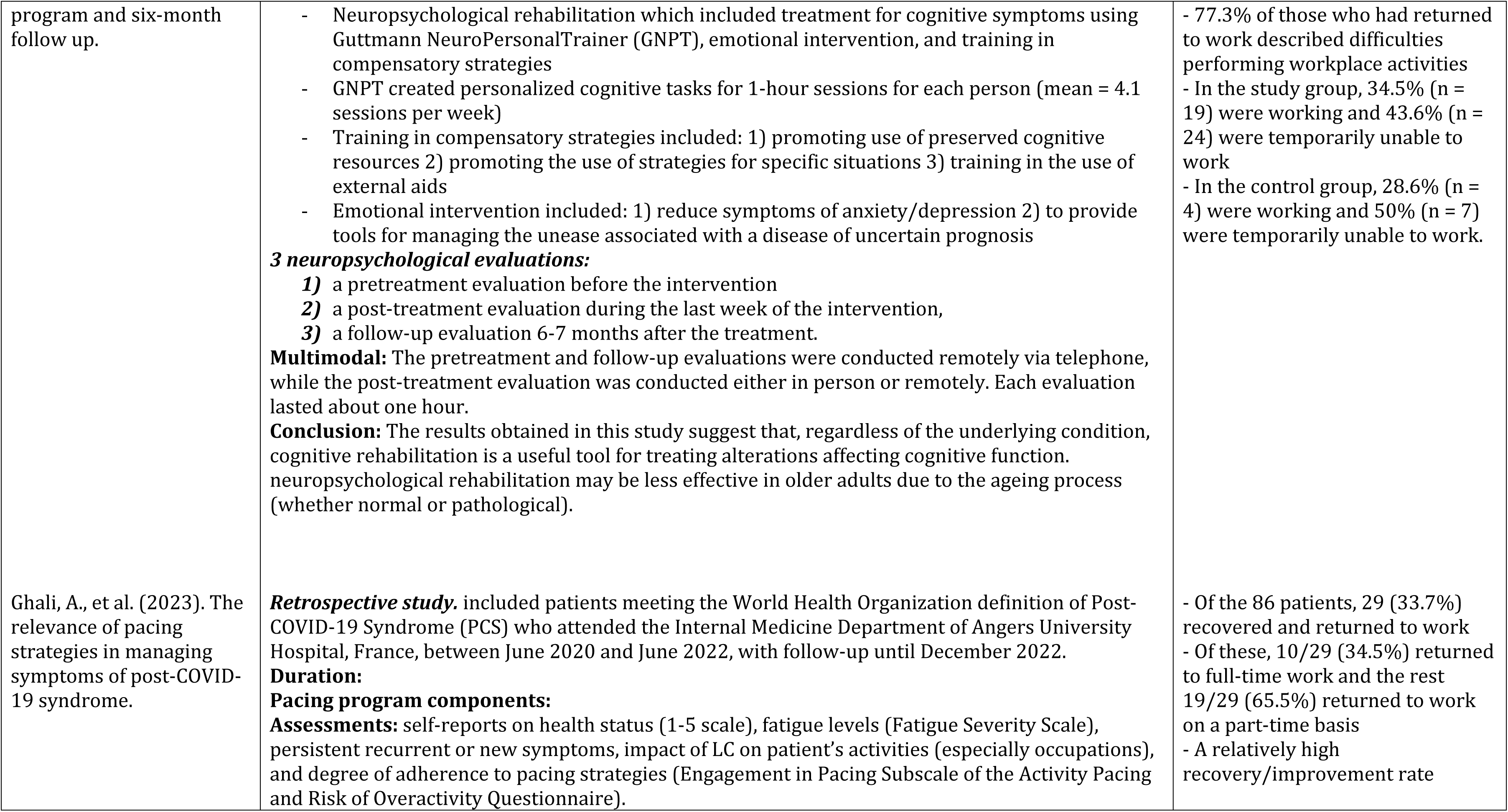
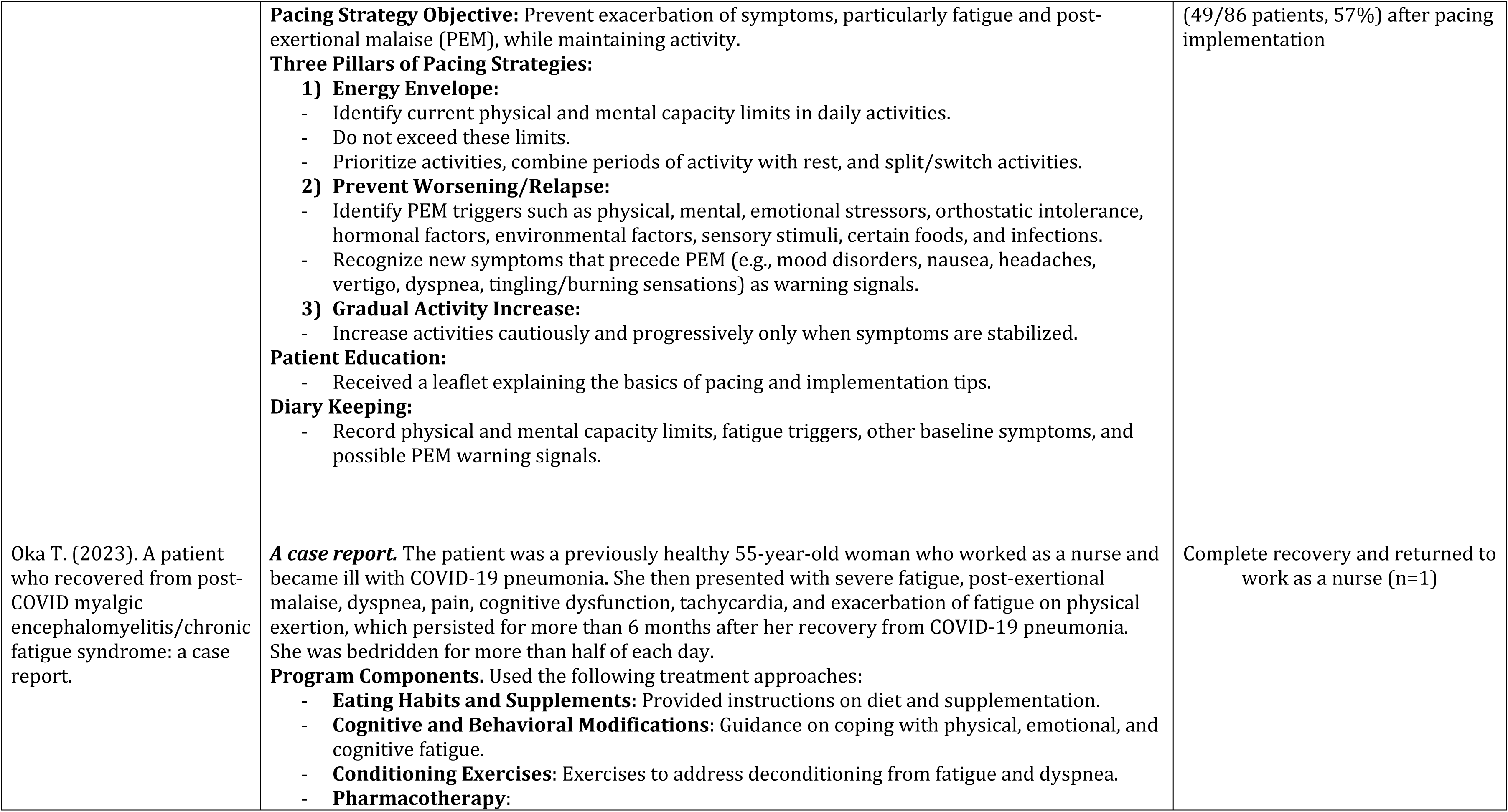
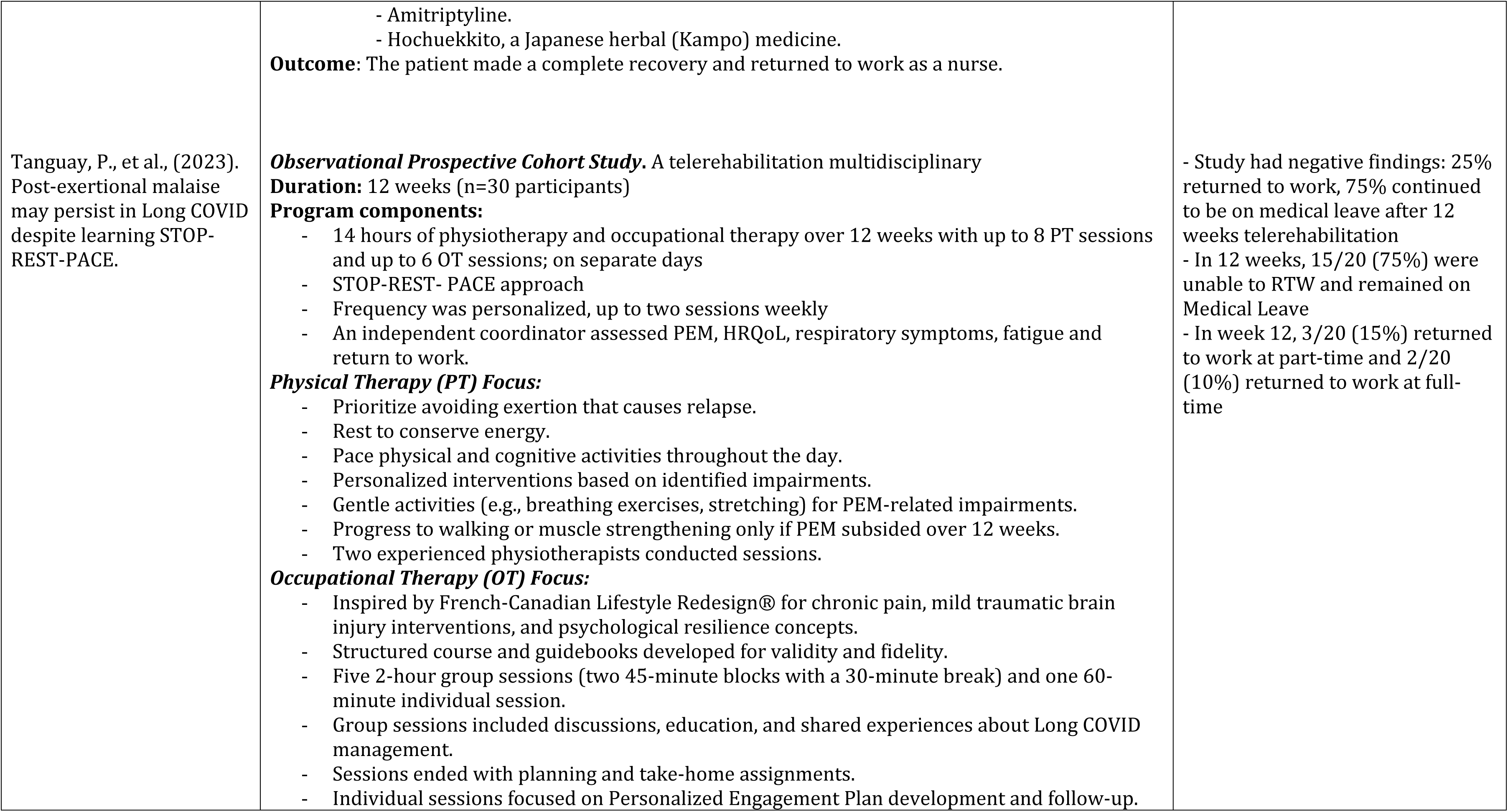
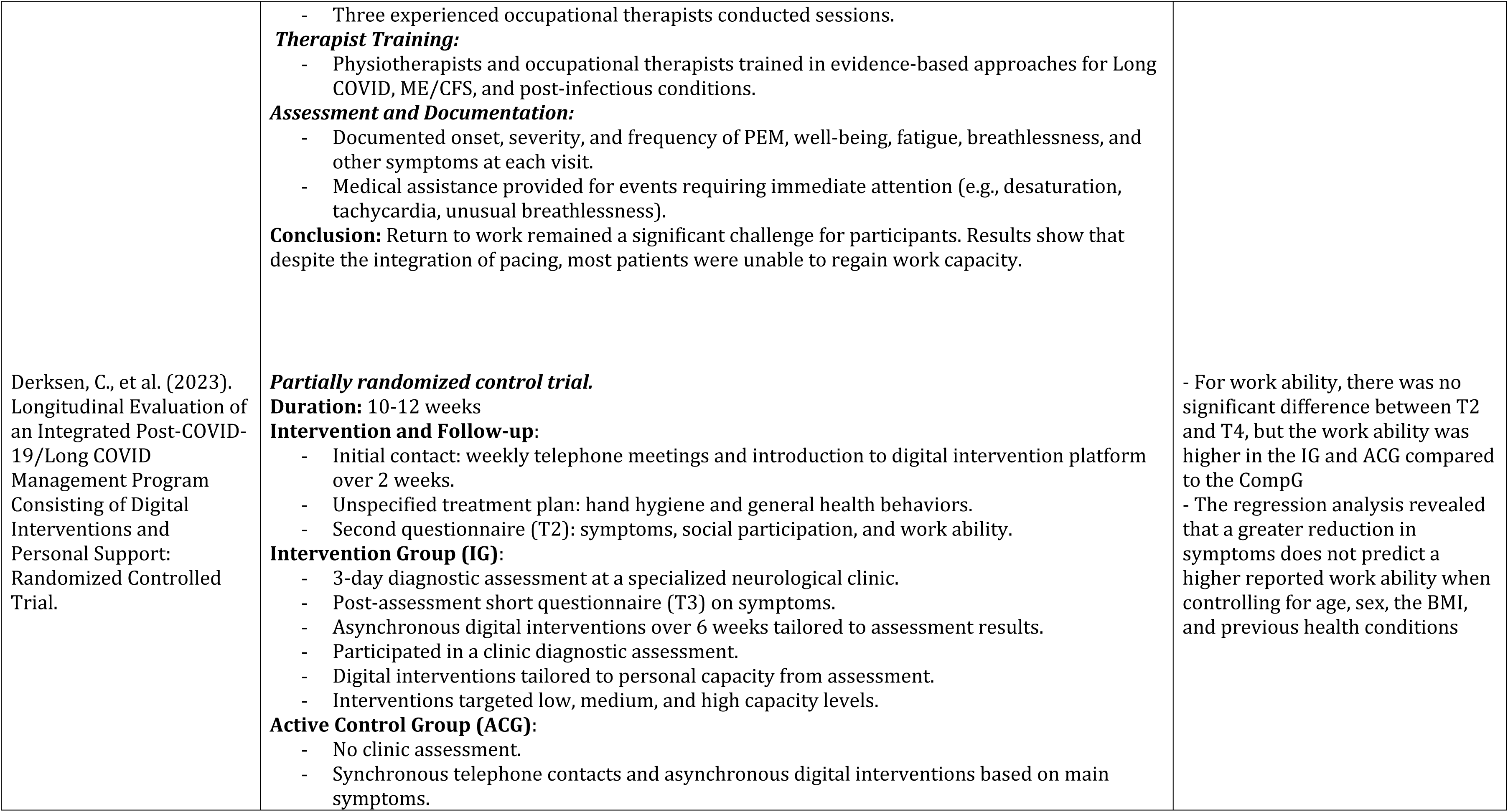
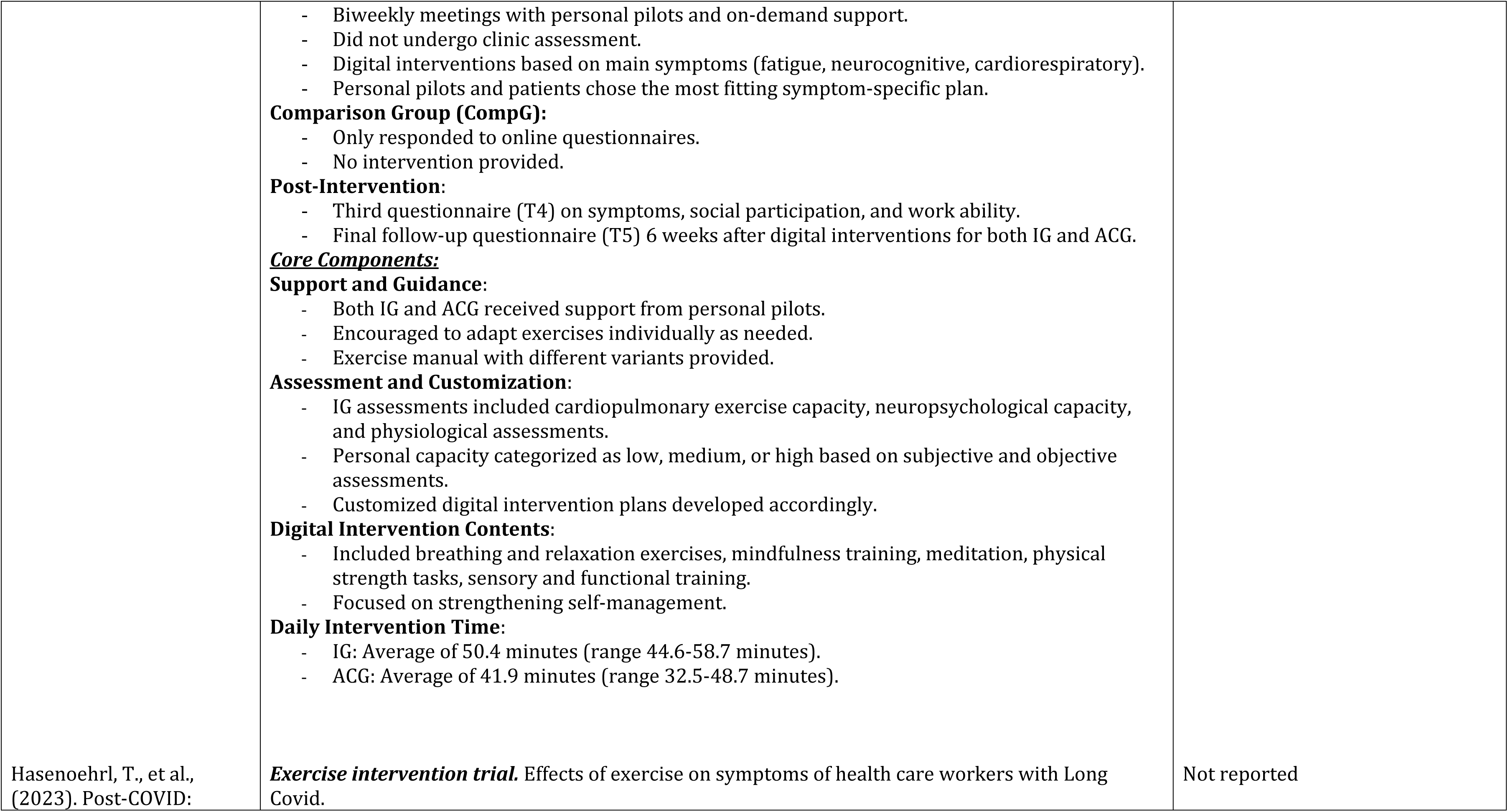
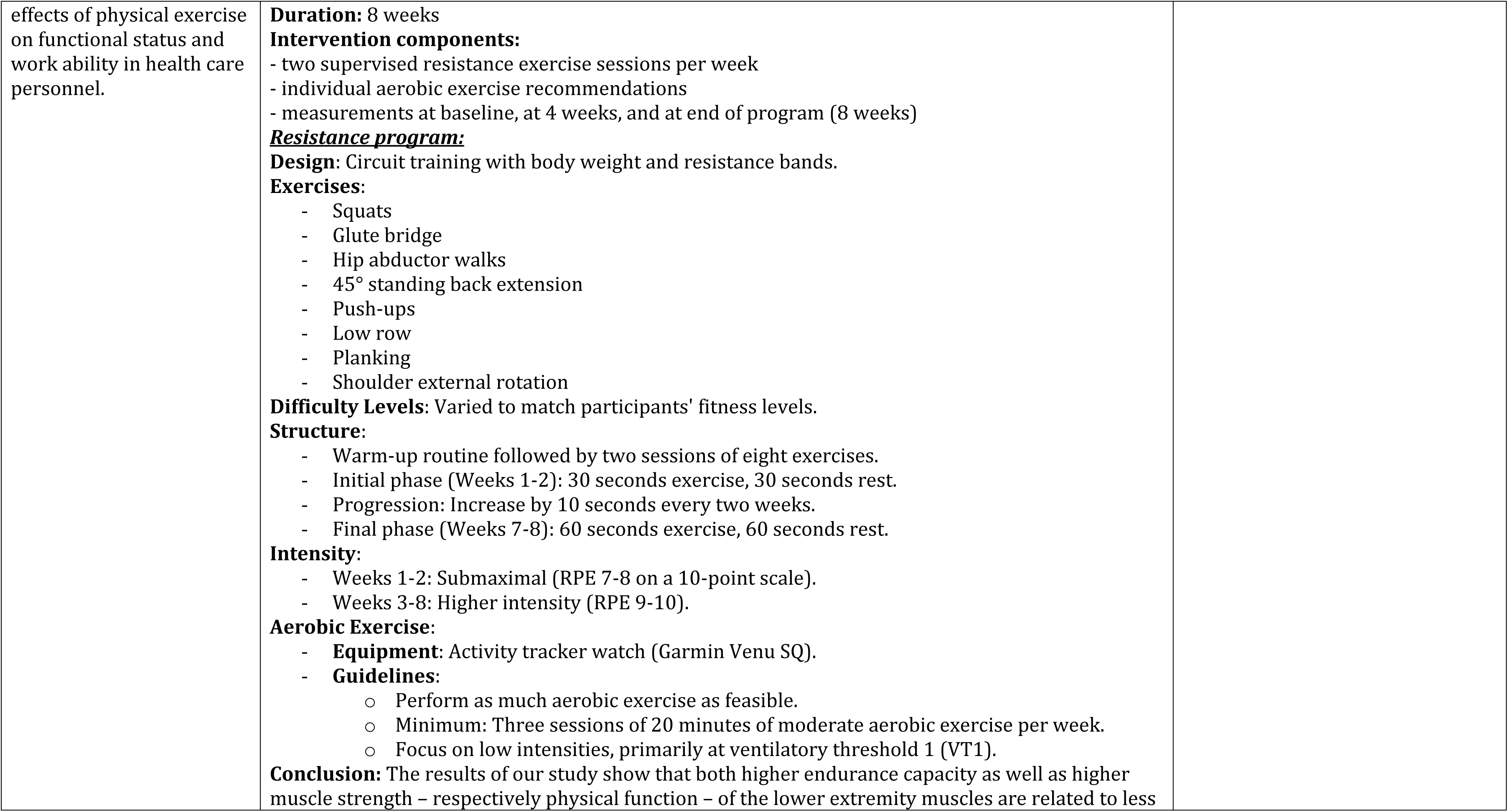
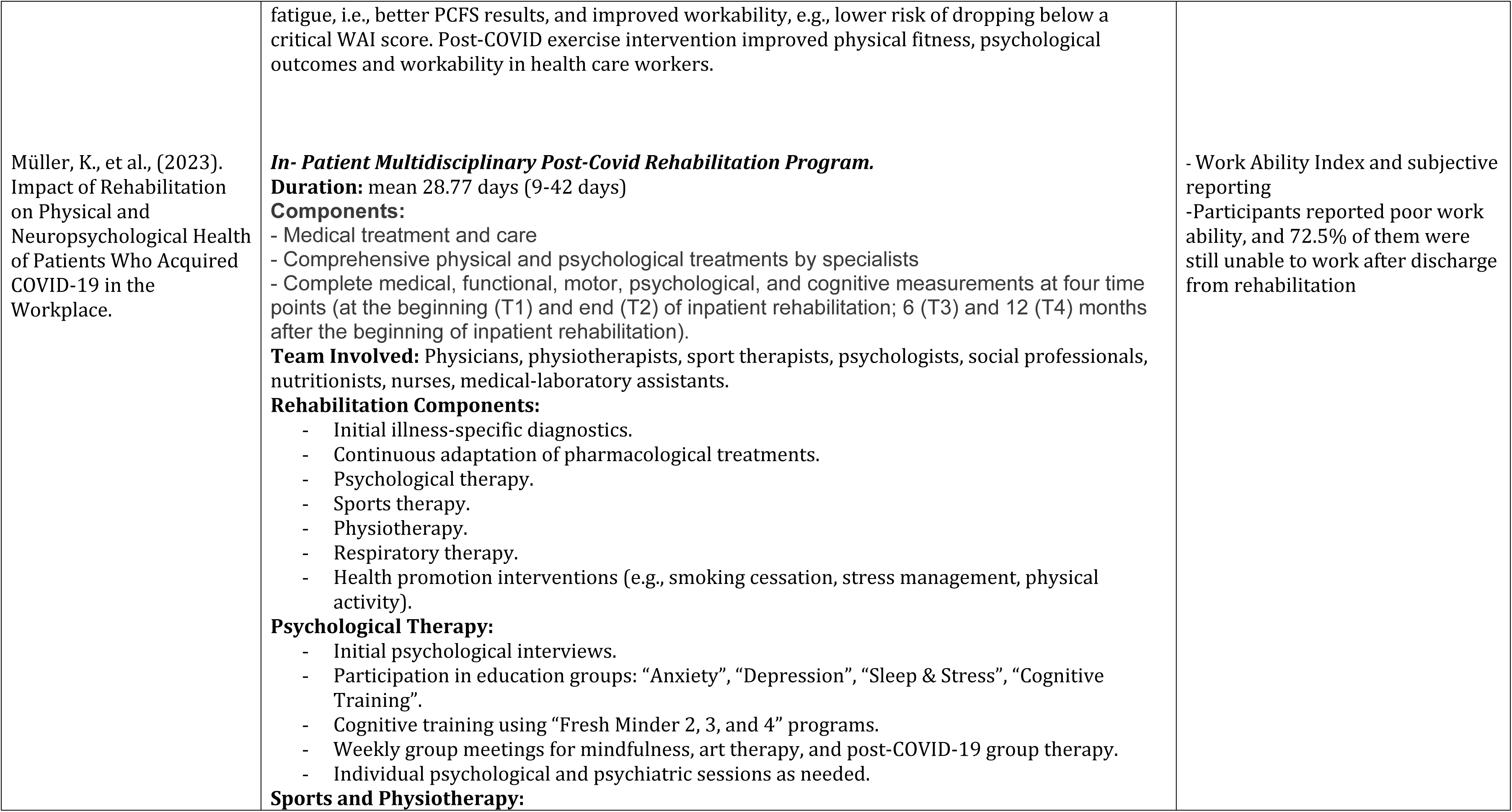
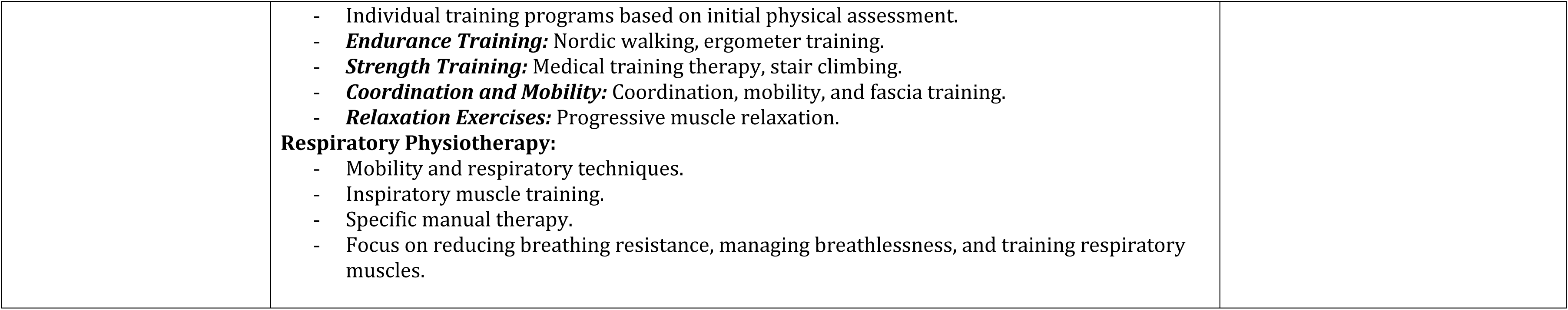
Categories and key messages of interventions studies for Long Covid.

**Table 3.**
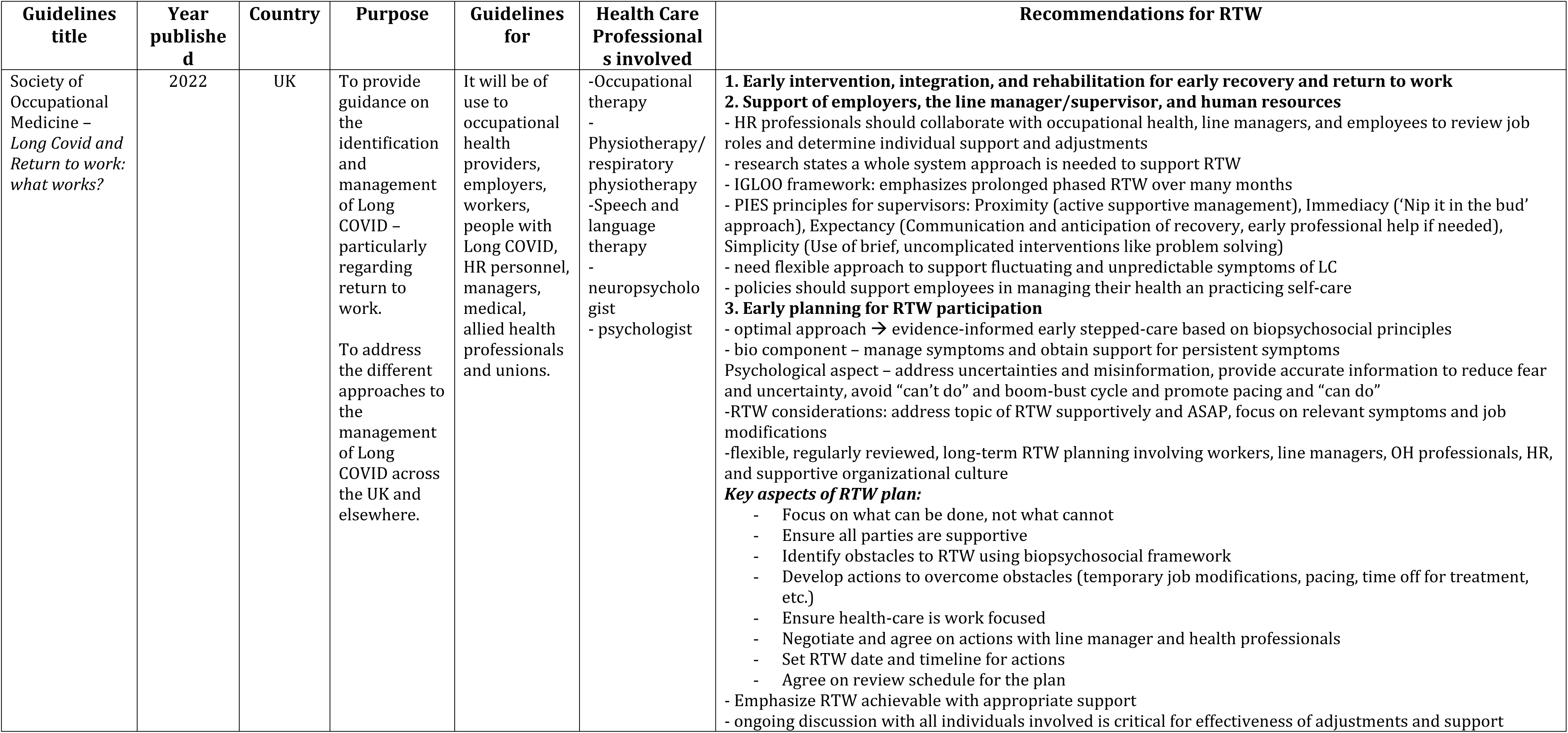
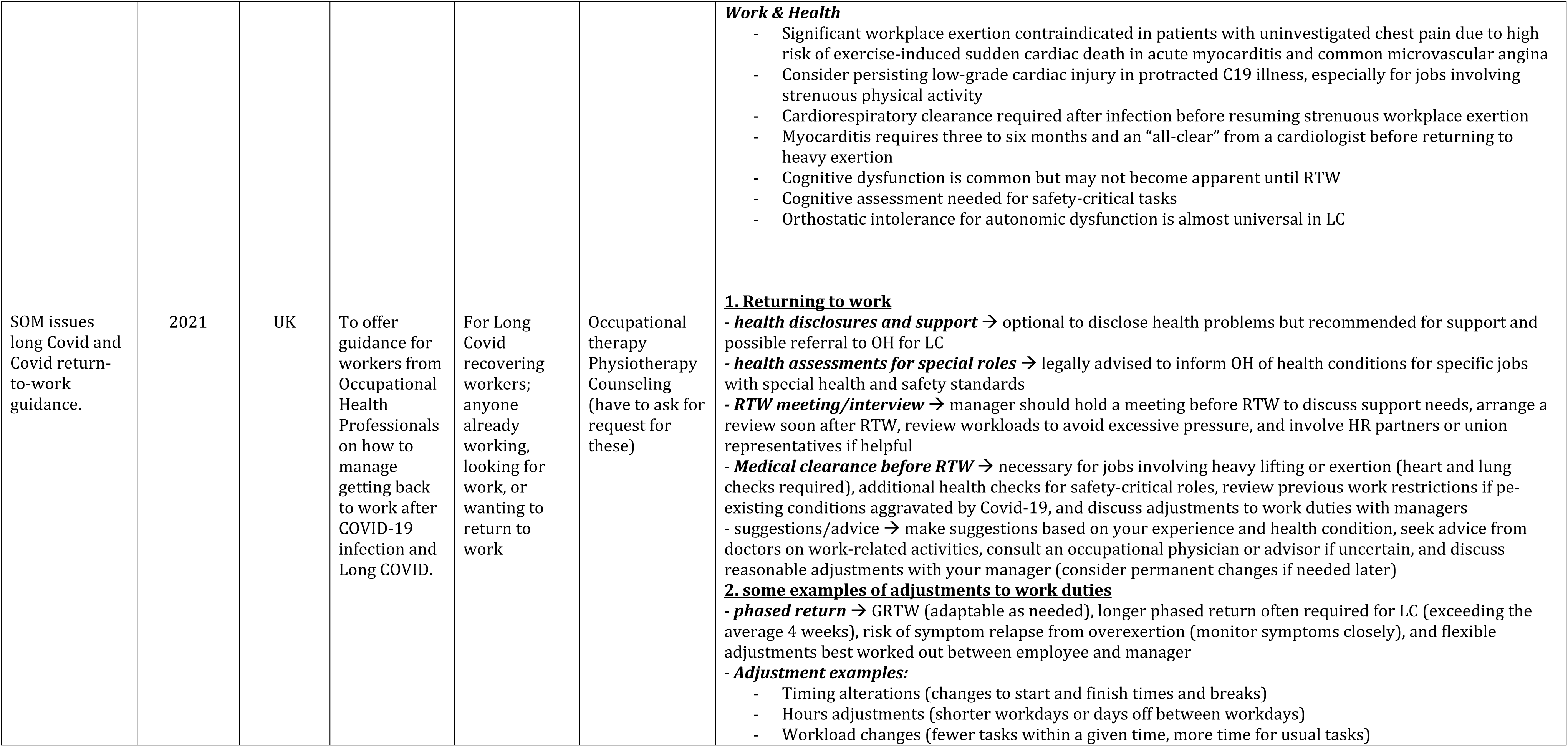
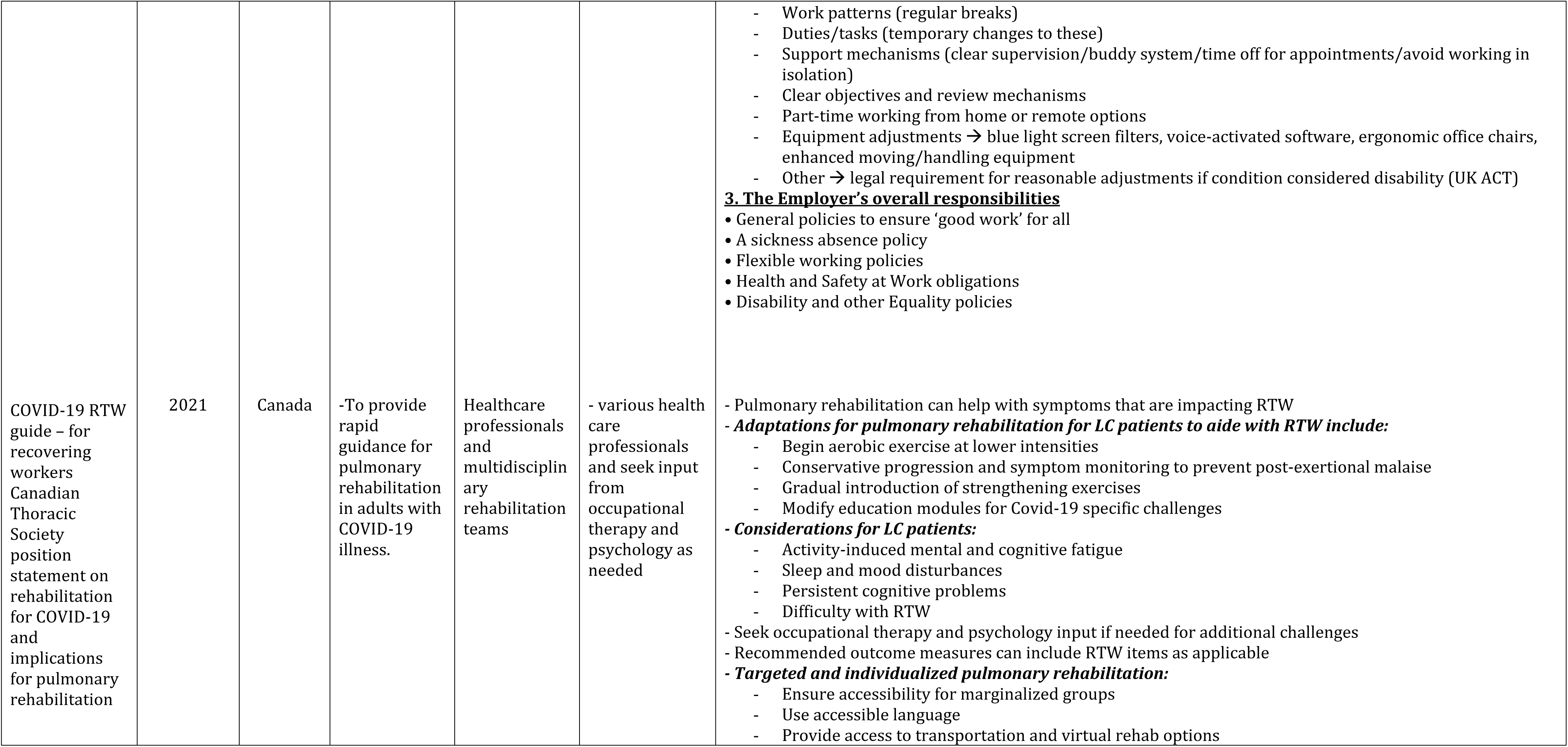
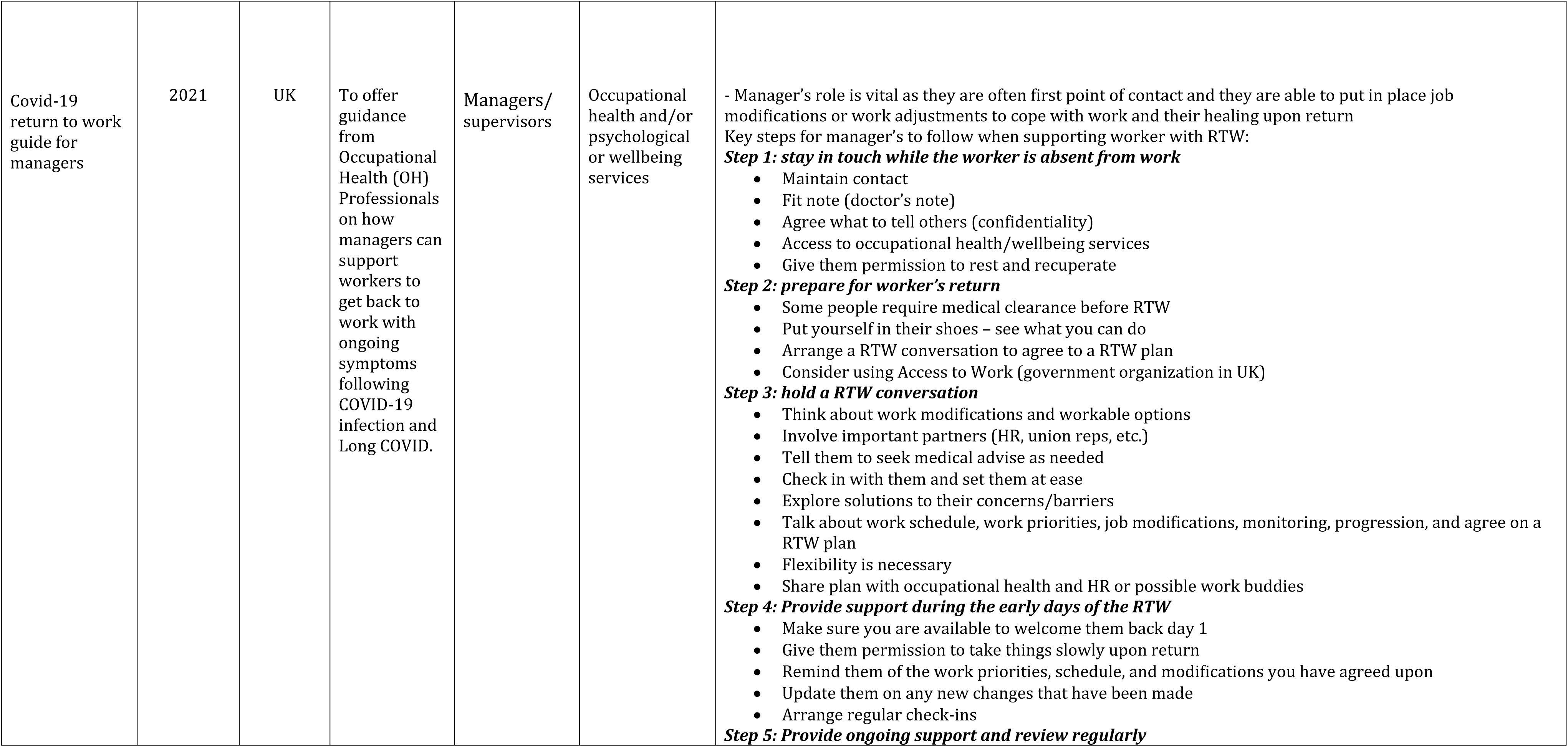
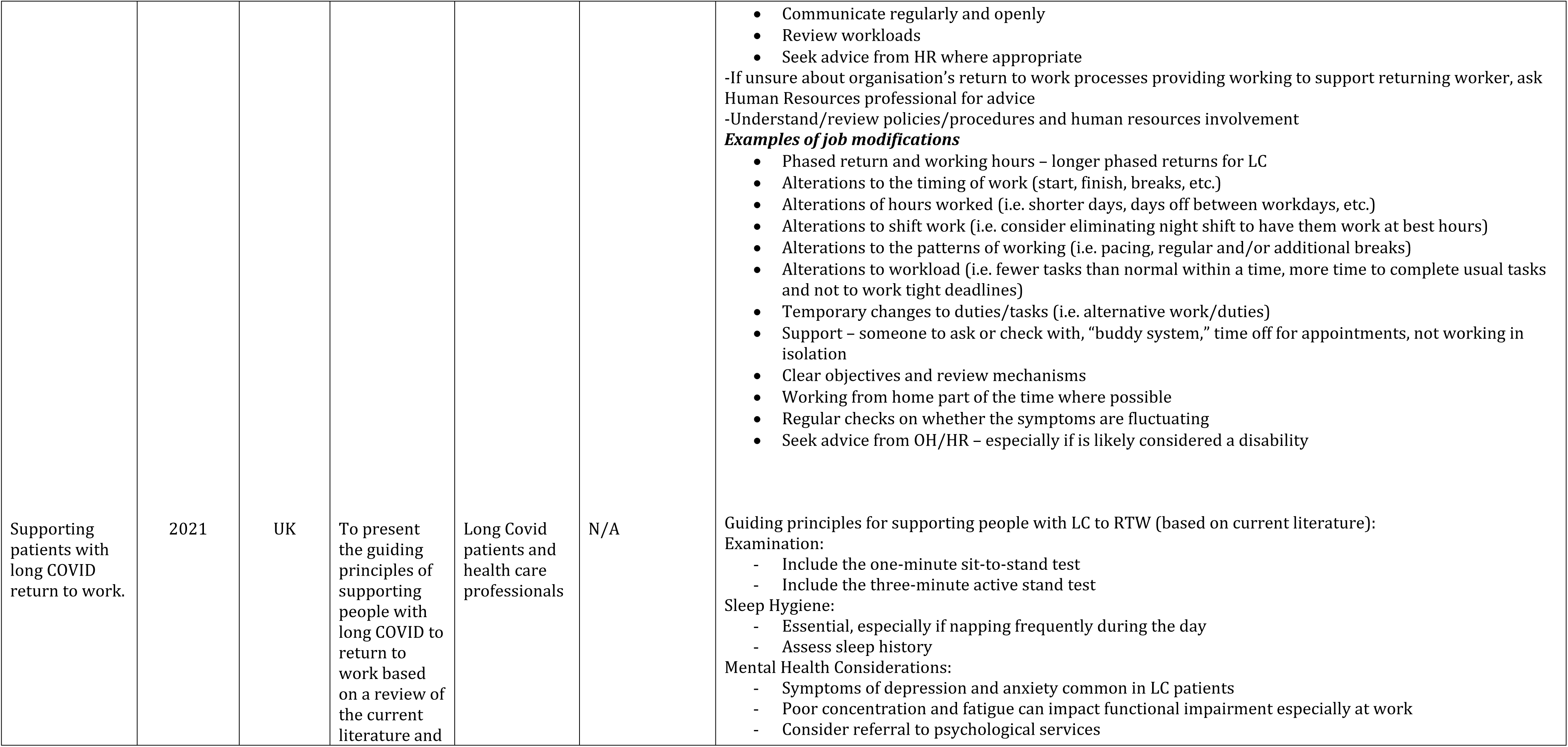
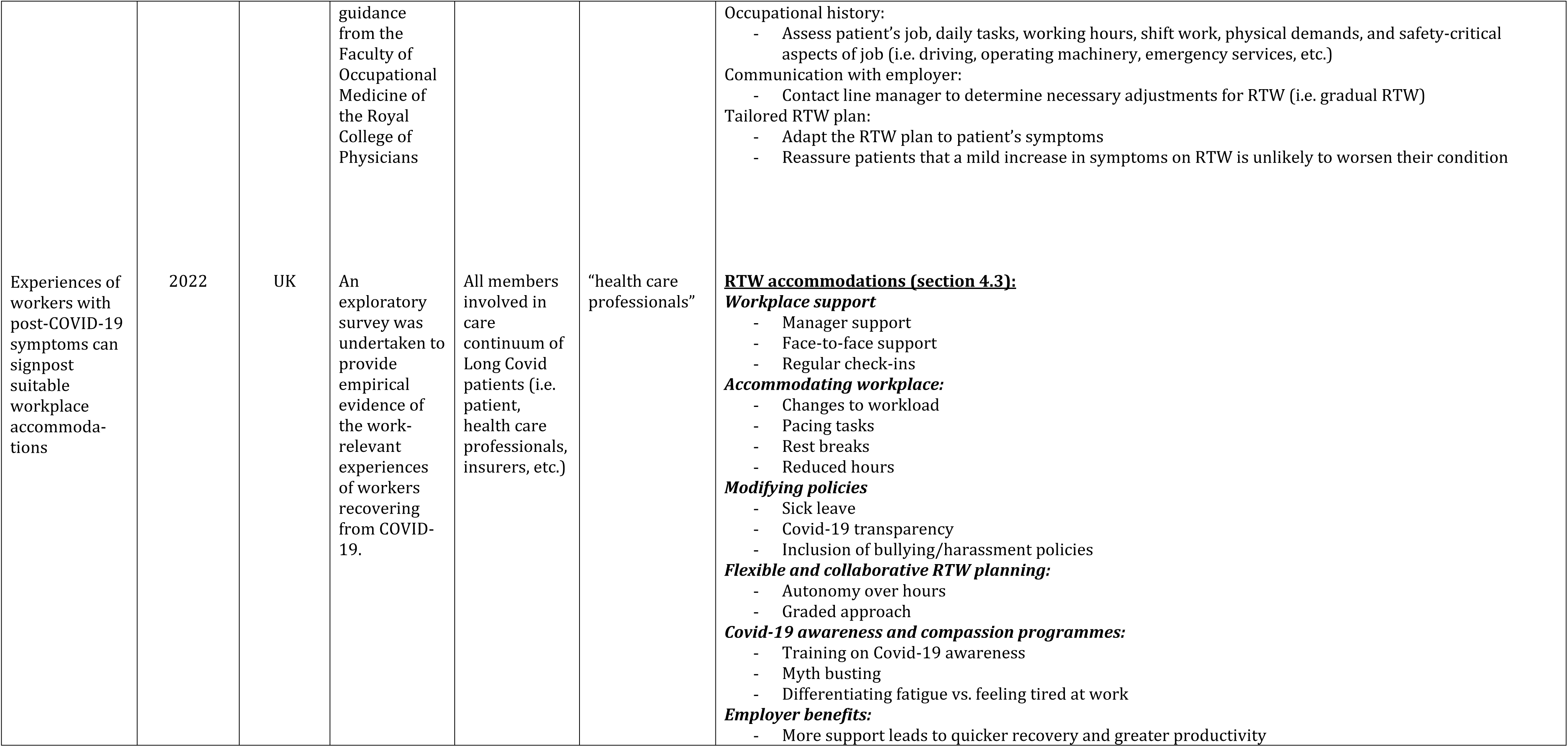
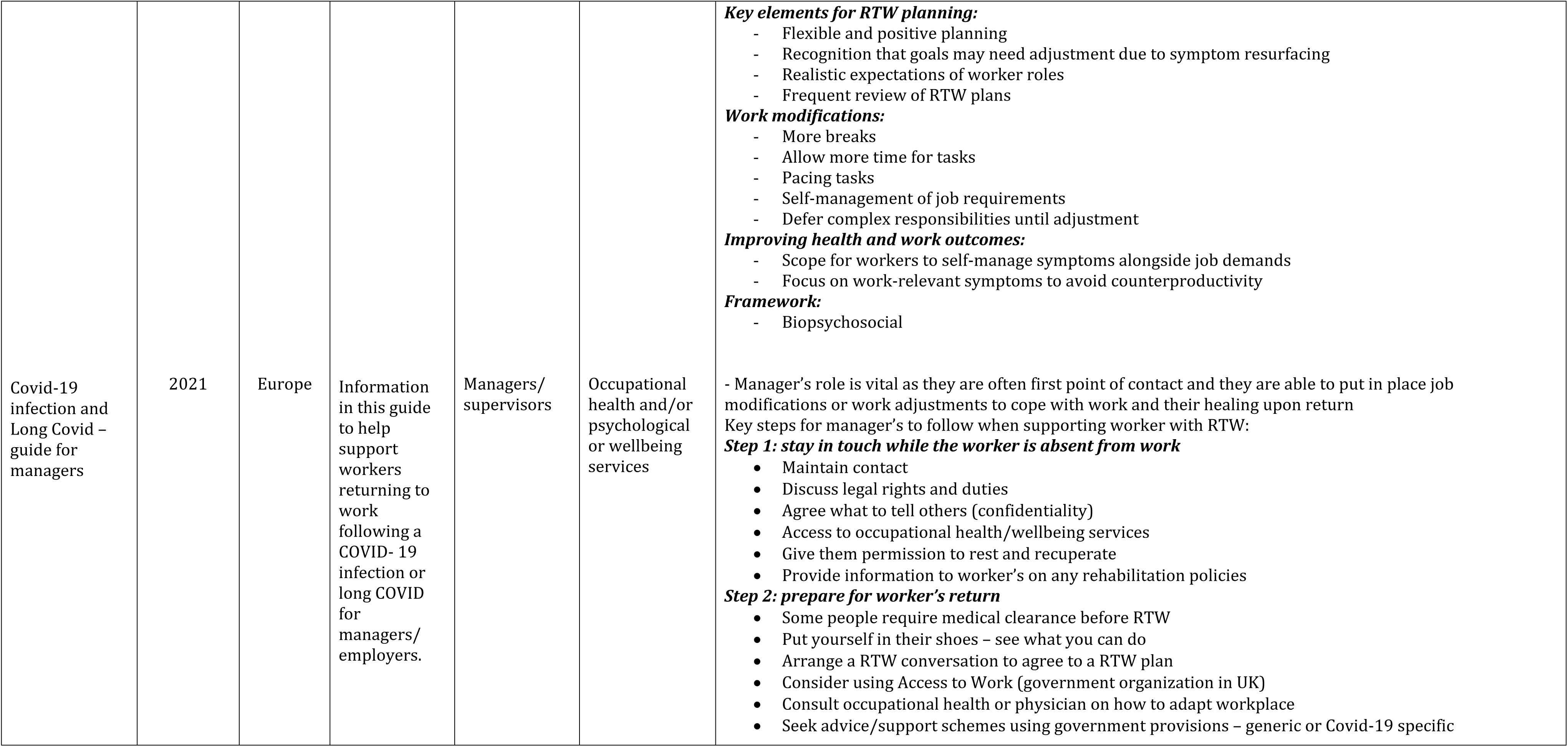
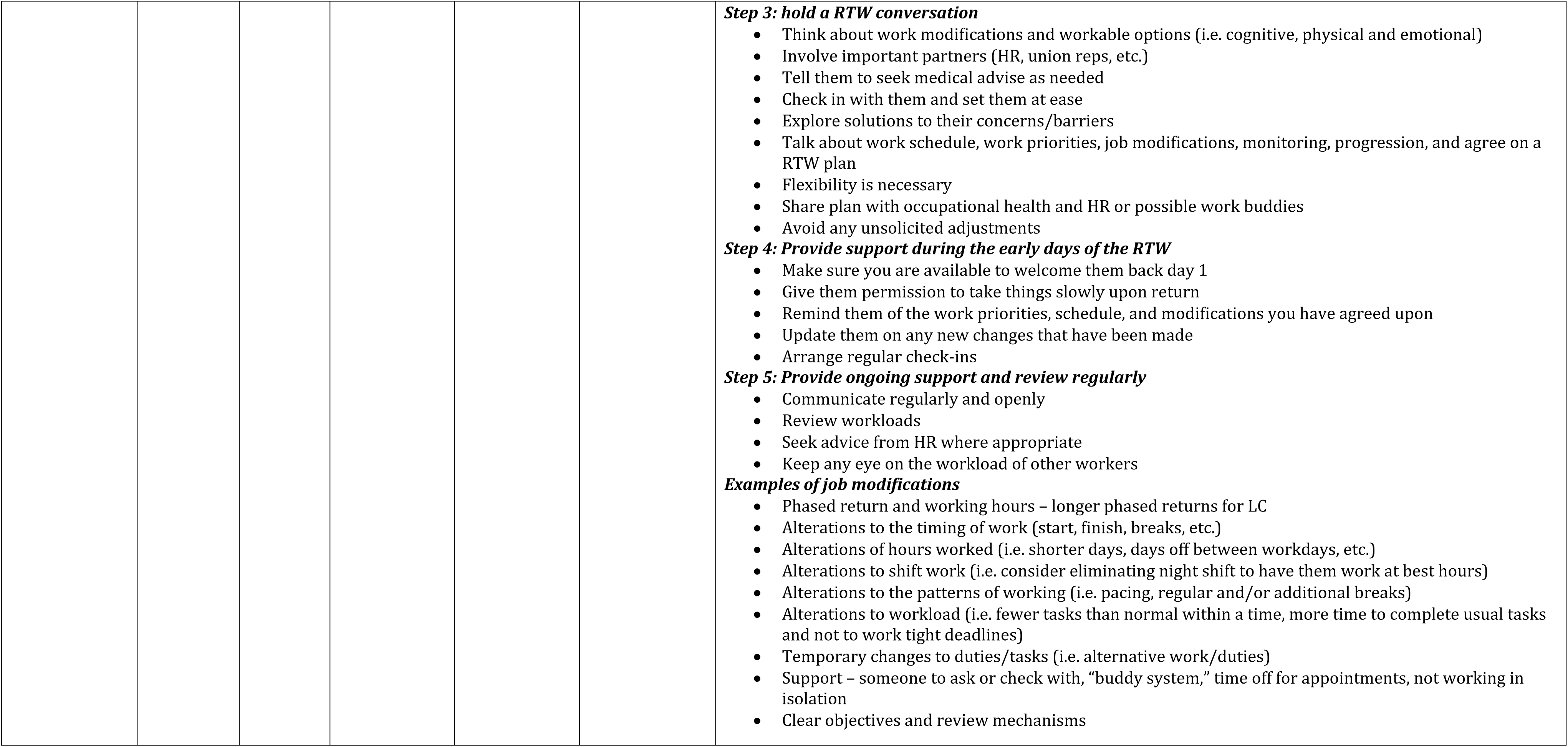
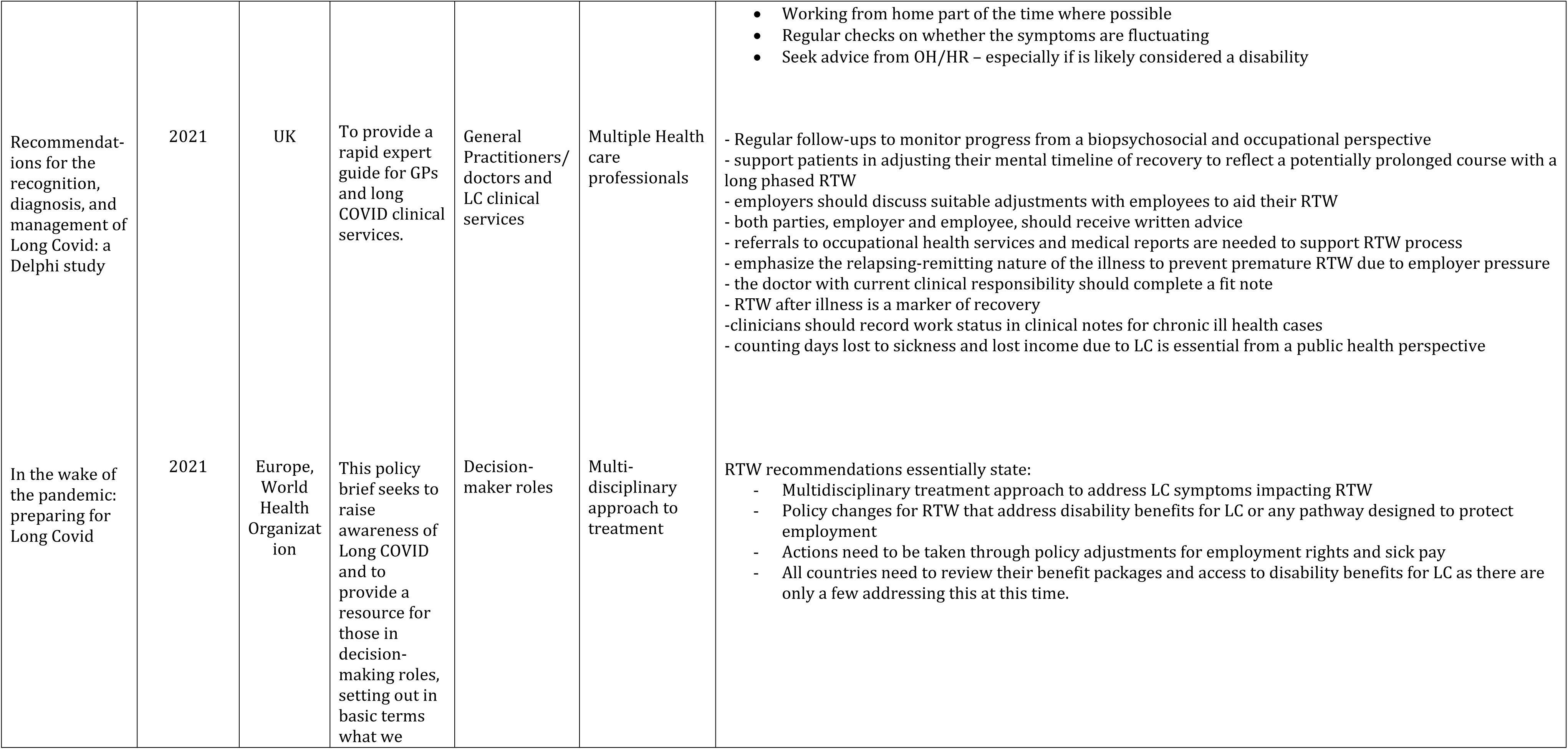
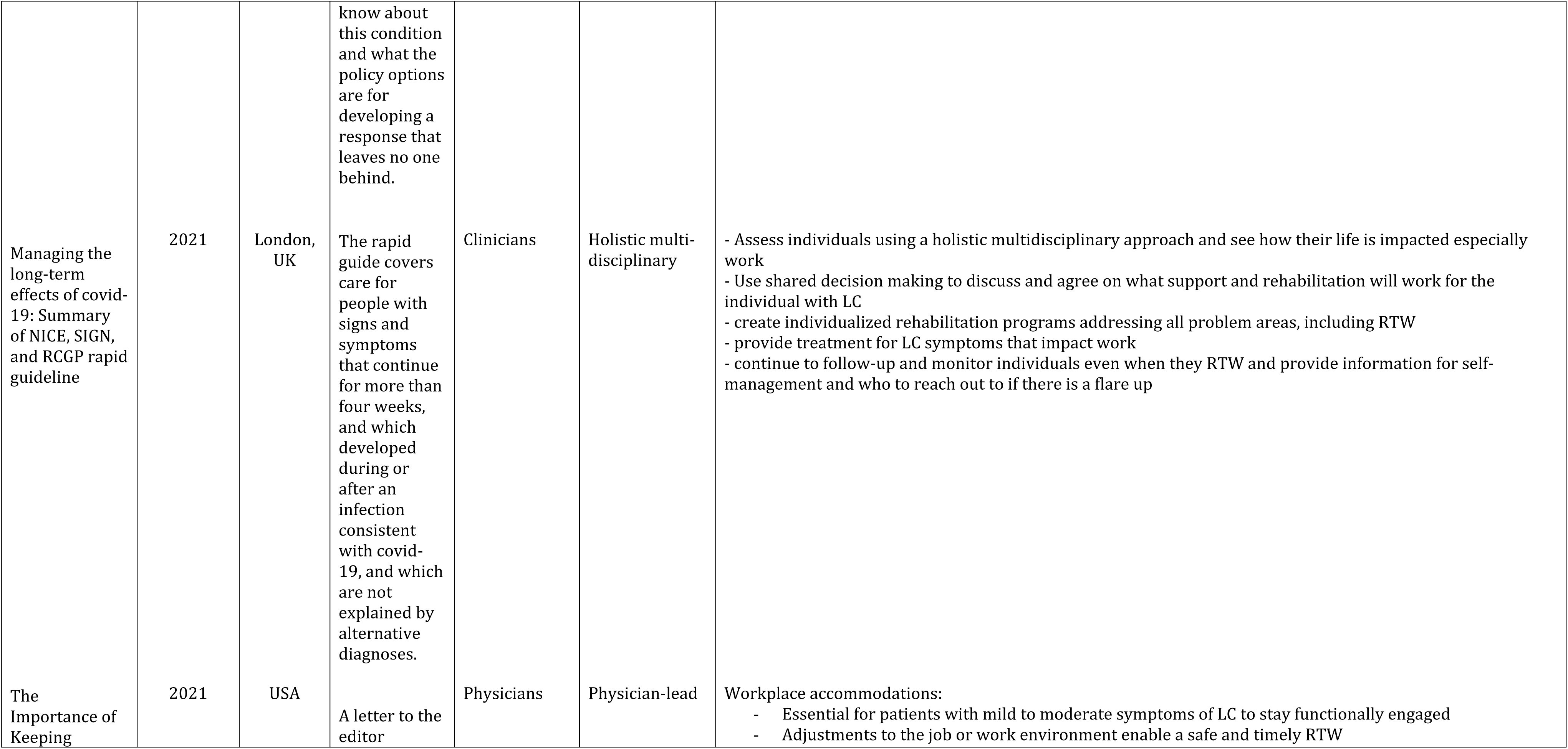
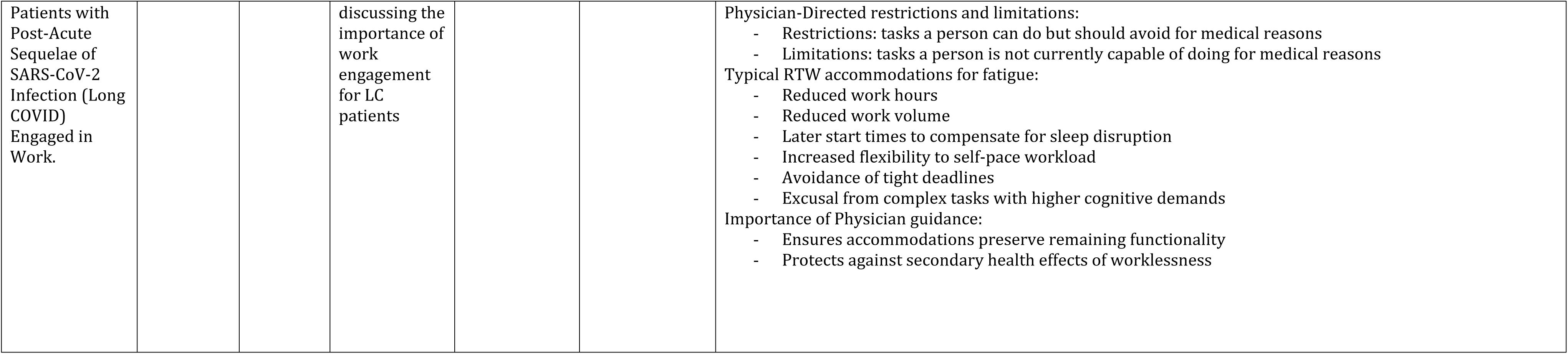
Recommendations for return-to-work from guidelines.

Relevant articles included a diverse range of study designs including three quasi-experimental studies (21–23), one randomized controlled trial (RCT) (24), six cohort studies (7, 15, 25–28), and one case report (29). We included three studies from Germany, one from Austria, one from Norway, one from Spain, one from France, two from Canada, one from the USA, and one from Japan. Notably, over half of the samples in all studies were composed of females, and the majority (63.6%) had a higher female sample.

Regarding the interventions, most lasted over three weeks (81.8%), with one study’s intervention lasting only three and a half days while the others ranged between nine and 42 days. All studies included participants over 18 years of age who met the Long COVID definition of either the World Health Organization or the National Institute for Health and Care Excellence (3, 30). Six studies required confirmed acute COVID-19 infection (21, 23–26, 29), while three allowed suspected or confirmed infection (26–28), and two had vague inclusion criteria without specifics related to COVID-19 (28, 31).

### 3.2. Key intervention findings

#### Effective strategies/approaches to treatment

The relevant studies highlighted several successful approaches to rehabilitation and treatment for managing Long COVID or RTW. These programs ranged from specialized, individualized, multiple-session rehabilitation to concentrated three-day rehabilitation programs.

##### Rehabilitation and Clinical Interventions – Physical Health

Rehabilitation programs that showed promising results for RTW included respiratory therapy, muscular training, a mental health component, and educational interventions (7, 21, 22). Two studies found that multidisciplinary, individualized treatment plans were recommended to address Long COVID symptoms like fatigue, autonomic dysfunction, and cognitive deficits, which impact workplace sustainability (7, 22). Altmann et al., (2023) found breathing exercises and respiratory treatments facilitated by physical therapists aided individuals in managing symptoms of shortness of breath and fatigue crucial for RTW (22). In addition, they found monitoring oxygen levels during rehabilitation and daily activities was helpful for pacing and managing PEM, contributing to sustainable RTW (22). It was found that RTW readiness from a physical stance, due to the variability of symptoms experienced, was challenging and required input from multiple experts and specialists (7).

##### Rehabilitation and Clinical Interventions – Mental Health

Altmann et al., (2023) also found that psychological counselling, using coping strategies such as mindfulness, helped manage anxiety and depression stemming from Long COVID itself, uncertainty about recovery, and grief over lost abilities (22). Early education on symptom management, pacing, graded return to activity, and energy conservation were emphasized as key components of interventions to manage Long COVID and build capacity for RTW (7, 26). Garcia-Molina et al., (2022) found that cognitive rehabilitation was effective for RTW by addressing cognitive impairments such as memory loss, brain fog, and concentration difficulties (23). Interventions like cognitive pacing, compensatory strategies, and memory exercises enhanced mental clarity and cognitive function, supporting RTW (23). In addition, Garcia-Molina et al., (2022) found that symptom monitoring tools like daily diaries or trackers helped individuals identify patterns in symptoms, enabling targeted interventions and a smoother RTW transition (23).

##### Workplace Strategies

Ongoing monitoring and individualized treatment plans at the workplace were considered promising with graduated RTW processes recommended to accommodate the fluctuating nature of Long COVID symptoms (7, 26). According to Brehon et al., (2022) and Garcia-Molina et al., (2022) RTW planning with modified, gradual returns (including adjusted hours and duties) was essential to accommodate fluctuating and episodic Long COVID symptoms and led to positive RTW outcomes. Frisk et al., (2023) adds another important component including RTW planning and suggestions for policy changes to sick leave to allow greater flexibility, remote work options, and time off for people living with Long COVID due to the episodic nature of symptoms (7, 21). These research studies found graduated RTW plans with modified work responsibilities were promising for RTW planning (7, 21, 23, 32). Early intervention (less than 6 months) was linked to higher RTW success rates, as shorter times between symptom onset and treatment correlated with improved outcomes via a higher likelihood of successful RTW (7).

##### Exploratory Medical Procedures

Other promising interventions included specialized rehabilitation programs for Long COVID such as using Enhanced External CounterPulsation (EECP) (22, 25). Sathyamoorthy et al., (2022) found that EECP improved blood flow and reduced cardiac strain, helping alleviate symptoms like persistent fatigue and shortness of breath, which in turn boosted physical endurance and overall well-being, facilitating a more sustainable RTW(25). The sample size for this study was small (n=16), thus this approach should be considered exploratory.

#### Approaches deemed less promising

Some interventions were found to have little impact on RTW in people living with Long COVID. Derksen et al. (2023) completed an intervention involving personal pilots and digital rehabilitation which was unsuccessful in improving RTW outcomes for people living with Long COVID, despite reductions in symptoms (24). Although personal pilots provided valuable support and helped patients navigate healthcare, their lack of therapeutic training and the reliance on asynchronous digital interventions without a physiotherapist’s supervision limited effectiveness (24). While symptom reduction predicted higher social participation, it did not translate into improved work ability, suggesting that work ability is influenced by broader social factors beyond individual symptom management (24).

Unsuccessful rehabilitation treatment options often involved aggressive or graded exercise plans without accounting for PEM, which worsened symptoms (28). Hasenoehrl et al.’s (2023) rehabilitation intervention focused on physical conditioning for individuals with Long COVID to improve endurance, muscular strength, and work ability (28). Although exercise led to significant improvements in physical fitness, psychological outcomes, and reported work ability, the individual gains were more pronounced in those with severe fatigue compared to those with milder symptoms (28). Despite these benefits, the intervention did not significantly enhance RTW overall suggesting that while exercise may lead to improvements in some outcomes, it does not address all the complex components of RTW, especially in individuals with varying levels of fatigue, PEM, and other Long COVID symptoms (28).

Muller et al. showed that rehabilitation targeting physical and neuropsychological health for individuals with Long COVID can lead to specific symptom reduction but not overall global fatigue (31). This study measured various aspects of each symptom across two time points with multiple tests and outcome measures, which documented a decrease of prevalence in all symptoms except for fatigue (31). Muller et al., found that despite the significant self-reported health improvements in people living with Long COVID, especially healthcare workers, they still continued to suffer from poor work ability with 72% of participants still unable to RTW post-rehabilitation (28, 31). Rehabilitation programs that lacked individualized treatment or had too many outcome measures were less effective and did not improve work ability (28, 31).

### 3.3. Recommendations related to RTW from published guidelines

Multiple guidelines related to management of Long COVID have been published in various countries and by the World Health Organization (30, 33–43). Most of these provide recommendations for RTW, which will be summarized here.

#### Individualized support

Guidelines for RTW recommendations emphasize tailored support acknowledging unique needs, symptoms, and personal situations as well as the importance of early intervention, integration of healthcare services, and personalized rehabilitation plans (33). Proactive involvement of individuals from various areas (clinical, workplace, unions, insurance/compensation, etc.) is important for finding solutions for successful RTW (33). Individualized, tailored treatment recommendations include starting aerobic exercises at lower intensities with conservative progression and careful symptom monitoring to avoid PEM (38, 44). In addition, it has been recommended that individual needs be recognized and addressed such as activity-induced mental and cognitive fatigue, sleep and mood disturbances, persistent cognitive problems, and other unique difficulties with RTW (38, 44). Adequate rest and medical assessment at four weeks post-symptom onset appears crucial for preventing Long COVID symptomology, followed by referrals to specialists and tailored rehabilitation programs as indicated (33). Treatment should focus on Long COVID symptoms that impact work readiness, with ongoing follow-up, symptom monitoring, and providing self-management resources and contact information for flare-ups (39, 42). Lastly, individualized accessibility recommendations ensure that specific therapies such as pulmonary rehabilitation are accessible to equity-deserving populations, use clear language for ease of interpretability, overcome transportation needs or provide virtual options to support participation in treatment (44).

#### Workplace accommodations

Workplace accommodations are crucial for people with mild to moderate Long COVID symptoms to stay engaged and safely RTW (39, 43). Recommendations for common accommodations for fatigue include reduced work hours, flexible scheduling, and avoidance of tasks with high cognitive demands (39, 43). Gradual RTW is often necessary due to the unpredictable nature of Long COVID symptoms (34, 36, 39). Work task or work environment modifications, guided by healthcare provider-directed restrictions and limitations, can often enable a safe and timely RTW (35, 40, 43). Physician and/or clinician guidance is essential to ensure that accommodations preserve functionality and protect against the negative health impacts of worklessness (40, 43). RTW planning meetings have been recommended prior to gradual RTW to address needs, review workloads, involve union and/or employer representatives and explore modifiable duties and responsibilities (35, 38, 39). Some modifications recommended include start and end time flexibility, modifiable hours/days, workload changes, increased breaks (i.e. microbreaks), modified or alternative duties, support mechanisms (i.e. a buddy or work from home options), and equipment adjustments (34–36, 39). Other recommendations include modifying policies (i.e. sick leave, bullying/harassment, confidentiality around sharing positive testing, etc.), fostering autonomy over scheduling (i.e. remote work, self-management of symptoms, etc.) and workplace goal setting (i.e. modifying targets) (39).

A key workplace accommodation recommendation for people living with Long COVID involves reviewing and changing policies that address disability benefits and employment protection pathways (39, 41). Actions should be taken to adjust policies concerning employment rights and sick pay as well as provide flexibility in leave due to the episodic nature of Long COVID (39, 41). It has been recommended that all countries review and update their wage replacement benefit packages and access to disability benefits for Long COVID (41).

#### Managers play a critical role

Managers play a significant role in facilitating a successful RTW as they are often the first point of contact for workers considering RTW after Long COVID (34, 36, 39, 40). RTW for people living with Long COVID requires support and collaboration among multiple parties, including the worker, employer, line manager, and healthcare professionals (33, 34, 36). Employers, Human Resources, and supervisors play key roles in providing flexible support, and implementing job modifications and work adjustments from gradual RTW plans to support recovery in the workplace (33, 34, 36). Supportive attitudes, active listening, and an accommodating nature is vital for RTW support (34, 36). In addition, recommendations for managers include staying in touch during absences from work, preparing for the worker’s return, conducting a RTW discussion, providing support during early RTW days, and implementing modifications (34, 36). Ongoing discussions and regular reviews are essential to address the episodic nature of Long COVID to ensure sustained work ability (33, 40). The main objective of discussions should be a gradual and adaptable RTW, alongside support for sustainable approaches to RTW, as employment is beneficial for health from the broader disability perspective as we need work and/or productivity for our overall health and the global economy (33, 39).

#### Clinical intervention or rehabilitation programs

For individuals with significant health concerns, such as chest pain or cognitive dysfunction, it is recommended to carry out specific assessments and clearances before resuming strenuous or safety-critical work (33, 40). Physical exercise as an intervention is contraindicated for those experiencing PEM and a fit note should be obtained prior to starting the program (33, 40). Strengthening exercises should be introduced gradually, and educational modules should be adapted for Long COVID-specific challenges and be introduced early (44). Occupational therapists and psychologists are recommended for Long COVID treatment in particular for RTW readiness (44). RTW interventions should always incorporate RTW-specific items in outcome measures as part of the assessment and follow-up to determine if the intervention was successful (40, 44).

An important recommendation from clinical practice guidelines related to RTW interventions is to use a holistic, multidisciplinary approach to assess how lives, especially work lives, have been impacted by Long COVID (42). RTW discussions in programs should involve occupational health services and health professionals to support recovery (33, 41, 42). Shared decision-making should guide the discussion with agreement on supports and rehabilitation strategies tailored to an individual’s needs (40, 42). Creating individualized rehabilitation programs that address all problem areas, including RTW, is crucial (42). It is recommended that interventions should be tailored to address sleep hygiene, mental health concerns such as depression, stress, and anxiety, assessment and treatment of occupational demands, and supervised physical exercise (38). Effective RTW planning should focus on overcoming obstacles using the biopsychosocial approach and maintaining an adaptable approach to support people living with Long COVID throughout their recovery (33, 39). This includes setting realistic expectations, focusing on what can be achieved, and addressing both physical and psychological aspects (33). At this time, guidelines state that initial examinations or assessments prior to interventions should include the one-minute sit-to-stand test and three-minute active stand test to screen for orthostatic intolerance and PEM (38).

## 4. DISCUSSION

Determining which Long COVID symptoms impact recovery and require rehabilitation are necessary for assessment and treatment purposes. Key strategies for treatment highlight effective RTW interventions combining physical and mental health support, including respiratory and muscular training, psychological counseling, cognitive rehabilitation, and symptom management tools (7, 21–23). The findings of this review underscore several critical insights into the current state of RTW interventions for individuals with Long COVID. There are three key findings of this review. First, there is a notable scarcity of rigorous studies specifically evaluating the effectiveness of RTW interventions tailored for Long COVID. This highlights the research gap which limits the evidence base needed to guide clinicians and policymakers in supporting individuals with Long COVID effectively. Secondly, valuable insights can be obtained from existing clinical practice guidelines, which offer a range of RTW recommendations. These guidelines, although not always validated specifically for Long COVID patient population, provide practical suggestions that can be applied and adapted to this emerging area of practice. The third key takeaway is the complexity of RTW for individuals with Long COVID, as the episodic and unpredictable nature of the condition presents unique challenges. Achieving successful RTW for these individuals typically demands sustained, adaptable support. This includes individualized, targeted healthcare and rehabilitation interventions to manage symptoms, workplace accommodations to address fluctuating abilities, and comprehensive social support systems. Workplace accommodations such as flexible hours and gradual RTW are recommended to address fluctuating symptoms of Long COVID and align with broader RTW recommendations for work disability cause by health conditions or illness (7, 30, 41, 42). Interventions with any success for RTW included RTW planning with modified duties and suggested policy updates for flexible sick leave.

RTW recommendations from international guidelines emphasize individualized, multidisciplinary support involving clinicians, employers, and policy updates to facilitate a sustainable RTW process (30, 34–36, 39). Key findings highlight employer support from supervisors or managers, gradual RTW, clinician or physician oversight of RTW plans, and changes to policies/procedures for flexibility to account for the episodic nature of Long COVID (33, 34, 36, 40, 43). RTW is currently viewed as a marker of a form of Long COVID recovery as most individuals with Long COVID struggle to RTW in any capacity (42).

### Interventions that should be avoided

#### Resistance training or aggressive exercise

Aggressive exercise such as resistance exercise training or graded activity, are often counterproductive for individuals with Long COVID due to the high prevalence of PEM and the episodic nature of fluctuating symptoms like fatigue, autonomic dysfunction, and cognitive impairments (3, 5, 14, 17, 28). Unlike traditional rehabilitation where exercise is used to improve functional abilities and work readiness, for individuals with Long COVID experiencing PEM, excessive physical exertion can exacerbate symptoms leading to prolonged recovery setbacks (3, 14, 17). Graded exercise for those with PEM is currently contraindicated and studies indicate that interventions ignoring PEM risk worsening fatigue and other symptoms, thus reducing the effectiveness of rehabilitation and could potentially delay RTW (3, 14, 17). Therefore, baseline measures and monitoring symptoms must be a component of treatment where physical conditioning is involved (26). Overall, the current recommendation is a conservative, individualized approach to physical activity that prioritizes pacing and symptom monitoring to ensure safety and RTW support for individuals living with Long COVID.

#### Digital or unsupervised treatment

Unsupervised or digital-only rehabilitation programs for Long COVID recovery may be insufficient, as they often lack personalized oversight necessary to manage complex, fluctuating symptoms effectively. Less effective treatment approaches, such as digital-only programs, often overlooked crucial elements such as pacing, orthostatic intolerance and PEM hindering RTW and/or led to no changes in global fatigue (24). For those with symptoms such as fatigue, brain fog, and autonomic dysfunction, supervision and modifications are frequently needed, which digital platforms alone often cannot support (3, 24, 28, 30). As such, programs that do not provide proper training for people with Long COVID or rely on digital interventions without supervision were found to be less effective (24). These treatment programs also have the potential to worsen PEM and impact overall recovery for individuals with Long COVID potentially delaying RTW (24, 28). Without direct feedback from clinicians, patients may overexert themselves or miss subtle warning signs of PEM, risking flare up hindering overall recovery and delaying RTW (14, 24). This indicates the need for a more comprehensive approach that is supervised, multidisciplinary, and individualized, targeting the individual’s personal and contextual needs. Virtual rehabilitation treatment with mindfulness, physical and sensory exercises but no clinician supervision was found to lead to symptom reduction but no RTW highlighting the need for clinician support as stated in other sources (7, 24, 28). Studies show that in-person, supervised care from clinicians is vital to ensure effective and safe pacing, modifications to program, and comprehensive support necessary for sustainable RTW (24, 45).

#### Excessive Outcome Measurements

Although Long COVID rehabilitation can address physical and psychological symptoms, programs addressing every symptom with multiple outcome measures may fall short in RTW preparation and managing fatigue (31). Using too many outcome measures or maximal testing with performance-based assessment of work ability may overwhelm patients and clinicians, detracting from effective rehabilitation. This could be due to lack of treatment options and over 200 possible symptoms for Long COVID being identified. Clinicians may feel pressured to document all problem areas. With too many measures, the attention on meaningful progress or change can be diluted, making it difficult to identify which interventions truly support Long COVID recovery for RTW. Individuals with Long COVID often struggle with global fatigue and PEM, which may add cognitive and physical strain on this population, further complicating the recovery process (14, 17). They often require pacing and energy conservation strategies to manage symptoms; thus, intensive programs with multiple outcome measures can be additionally fatiguing for these individuals’ exacerbating PEM or worsening symptoms (26). There is a need to streamline outcome metrics to focus on core areas such as functional capacity, symptom management, and RTW readiness to allow clarity for more actionable insights, helping both clients and clinicians with targeted support that aligns with the unique and variable needs of each person with Long COVID.

### Limitations

This study included mainly English-language studies, which may have missed some important literature published in other languages. We included a very broad range of interventions, which included some exploratory medical interventions such as EECP. While interesting, these were not comparable to other commonly used RTW interventions and should be evaluated and assessed separately. The dynamic and rapidly-developing nature of this literature will require frequent updates.

### Recommendations for future research

Despite current rehabilitation recommendations and research studies for Long COVID, further research is necessary to refine rehabilitation programs to improve work ability and facilitate successful RTW (7, 21, 26). Further research is necessary to determine whether Long COVID is a single condition or a group of conditions requiring distinct treatment and monitoring (3). This can inform which professionals are necessary on multidisciplinary or transdisciplinary teams (20, 25). It is inevitable that Long COVID, like other chronic conditions, will require ongoing management due to its multidimensional, episodic, and fluctuating nature (13). Thus, research to determine similarities and differences between Long COVID trajectories and other chronic conditions may help inform Long COVID treatment approaches (3, 26). Individuals with Long COVID often face a minimum 3-month waiting period before accessing treatment such as rehabilitation programs, which may be a barrier to RTW (16). Thus, research on whether earlier education or access to rehabilitation interventions benefit individuals with lingering COVID-19, but not yet diagnosed with Long COVID, is necessary (16).

Future research should explore adapting rehabilitation programs for chronic illnesses for those living with Long COVID to improve work ability outcomes (7, 31). Much like other chronic illnesses, early rehabilitation for RTW needs to be explored for individuals with Long COVID as research findings correlate with RTW improvements for those with less time between symptom onset and rehabilitation (7). It has been highlighted that flexible work arrangements improve RTW for individuals with Long COVID thus, this should be incorporated into rehabilitation programs and tested in future studies (7). Additionally, personalized rehabilitation approaches incorporating work simulation of specific job demands and managing the severity of fatigue should be explored with education on pacing included (26). The benefits of early education on pacing, energy conservation, and compensatory cognitive strategies need to be tested in future research studies as individuals with Long COVID and current research point towards potential success for RTW in these areas (7, 23). Research on indicators for successful RTW and prognostic factors is also needed.

Exploring discrepancies across Long COVID rehabilitation programs and approaches is also necessary to determine their true correlations with RTW success. Specifically, the requirement for medical clearance or fit notes varies by employer, with some programs deeming RTW as a sign of symptom recovery/improvement while others view it as part of an ongoing RTW journey (7, 25, 26). Furthermore, variations in communication approaches regarding Long COVID affect the certainty of findings. Some programs prioritize autonomy and discretion, while others encourage openness about COVID-19 symptoms and infection. This is vital in the broader scope of Long COVID rehabilitation where agreement in diagnosis, definition, and prognosis across clinical team members is necessary (3). Understanding these discrepancies in current rehabilitation approaches can lead to standardization of best practices and improve rehabilitation outcomes for RTW in Long COVID (24, 26).

Despite the availability of numerous rehabilitative approaches to RTW for Long COVID available, specific rehabilitative treatment modalities and approaches for Long COVID rehabilitation are still necessary. To date, most approaches have considered a trial-and-error approach by applying strategies used with other populations, using traditional rehabilitation strategies, and/or using treatments that target individual symptoms; however, specific Long COVID treatment is necessary (24, 26). Longitudinal studies are crucial in understanding the sustainability of rehabilitation for RTW alongside ongoing aftercare support due to the episodic nature and fluctuations of Long COVID (7). Most programs have incorporated a mental health component, but further research is necessary to explore the psychological aspects of work readiness to develop strategies to enhance mental resilience optimizing RTW outcomes for individuals with Long COVID (21, 22). Health systems and policy research is also needed on the effect of disability benefits (short term or long term) and their effect on RTW.

## 5. CONCLUSION

Our findings highlight the importance of targeted, individualized rehabilitation approaches for individuals with Long COVID to support their recovery and facilitate sustainable RTW. There is a scarcity of rehabilitation tools or protocols specifically designed for this population to address Long COVID symptomology. This review highlights key elements of effective interventions to date such as integrating physical and mental health support, tailored and supervised pacing, and flexible workplace accommodations that align with the fluctuating nature of Long COVID symptoms. While some existing strategies have shown promise, other interventions do not appear to provide adequate support to promote RTW. Findings of this review highlight the necessity of refining rehabilitation programs in order to provide multidimensional, patient-centered, safe, and effective approaches to promoting RTW for this population. There is an urgent need to develop effective programs for individuals with Long COVID to promote recovery and RTW, especially given the social and economic impact of this condition and the current lack of appropriate RTW supports.

## Data Availability

All data use in this systematic review are publicly available.

## Funding

Funding for this research was provided by the Glenrose Rehabilitation Hospital Foundation.

## 6. AAPPENDICES

(See attached Appendix A)

1 figure see attached

## Notes

### Competing Interest Statement

The authors have declared no competing interest.

## REFERENCES

1. Merad M, Blish CA, Sallusto F, Iwasaki A. The immunology and immunopathology of COVID-19. Science. 2022;375(6585):1122-lp7.

2. Canada Go. Post-COVID-19 Condition in Canada: What We Know, What We Don’t Know and a Framework for Action.; 2022 December 2022.

3. Canada Go. TaskForce on Post-COVID-19 Condition in Canada: What we know, what we don’t know, and a framework for action.

4. Dennis A, Wamil M, Alberts J, Oben J, Cuthbertson DJ, Wootton D, et al. Multiorgan impairment in low-risk individuals with post-COVID-19 syndrome: a prospective, community-based study. BMJ Open. 2021;11(3):e048391.

5. Tabacof L, Tosto-Mancuso J, Wood J, Cortes M, Kontorovich A, McCarthy D, et al. Post-acute COVID-19 Syndrome Negatively Impacts Physical Function, Cognitive Function, Health-Related Quality of Life, and Participation. Am J Phys Med Rehabil. 2022;101(1):48–52.

6. Inclusively. New Research: The Immense Impact of Long COVID on Workers (And What Employers Can Do About It). December 2022.

7. Brehon K, Niemelainen R, Hall M, Bostick GP, Brown CA, Wieler M, et al. Return-to-Work Following Occupational Rehabilitation for Long COVID: Descriptive Cohort Study. JMIR Rehabil Assist Technol. 2022;9(3):e39883.

8. Brehon K, Miciak M, Hung P, Chen SP, Perreault K, Hudon A, et al. “None of us are lying”: an interpretive description of the search for legitimacy and the journey to access quality health services by individuals living with Long COVID. BMC Health Serv Res. 2023;23(1):1396.

9. Anderson E, Hunt K, Wild C, Nettleton S, Ziebland S, MacLean A. Episodic disability and adjustments for work: the ‘rehabilitative work’ of returning to employment with Long Covid. Disability & Society. 2024:1–23.

10. Canada Go. <COVID-19-longer-term-symptoms-among-Canadian-adults--Fourth-Report.pdf>. 2024 August 21, 2024.

11. Quinn KL, Lam GY, Walsh JF, Bhereur A, Brown AD, Chow CW, et al. Cardiovascular Considerations in the Management of People With Suspected Long COVID. Can J Cardiol. 2023;39(6):741–53.

12. Choutka J, Jansari V, Hornig M, Iwasaki A. Unexplained post-acute infection syndromes. Nat Med. 2022;28(5):911–23.

13. Davis HE, McCorkell L, Vogel JM, Topol EJ. Long COVID: major findings, mechanisms and recommendations. Nat Rev Microbiol. 2023;21(3):133–46.

14. Vink M, Vink-Niese A. The Updated NICE Guidance Exposed the Serious Flaws in CBT and Graded Exercise Therapy Trials for ME/CFS. Healthcare (Basel). 2022;10(5).

15. Muller K, Zwingmann K, Auerswald T, Berger I, Thomas A, Schultz AL, et al. Rehabilitation and Return-to-Work of Patients Acquiring COVID-19 in the Workplace: A Study Protocol for an Observational Cohort Study. Front Rehabil Sci. 2021;2:754468.

16. Pouliopoulou DV, Macdermid JC, Saunders E, Peters S, Brunton L, Miller E, et al. Rehabilitation Interventions for Physical Capacity and Quality of Life in Adults With Post-COVID-19 Condition: A Systematic Review and Meta-Analysis. JAMA Netw Open. 2023;6(9):e2333838.

17. Thille P, Cooper JE, Jansson A, Leclair L, Parsons J, Webber S, et al. Advocating for Long COVID Rehabilitation Support in Manitoba: An Environmental Scan.: Manitoba Academic Rehabilitation Sciences Covid Interest Group (MARSCI); 2022 2022-01-31.

18. von Zweck C, Naidoo D, Govender P, Ledgerd R. Current Practice in Occupational Therapy for COVID-19 and Post-COVID-19 Conditions. Occup Ther Int. 2023;2023:5886581.

19. Manhas KP, O’Connell P, Krysa J, Henderson I, Ho C, Papathanassoglou E. Development of a Novel Care Rehabilitation Pathway for Post-COVID Conditions (Long COVID) in a Provincial Health System in Alberta, Canada. Phys Ther. 2022;102(9).

20. Levac D, Colquhoun H, O’Brien KK. Scoping studies: advancing the methodology. Implement Sci. 2010;5:69.

21. Frisk B, Jurgensen M, Espehaug B, Njoten KL, Softeland E, Aarli BB, et al. A safe and effective micro-choice based rehabilitation for patients with long COVID: results from a quasi-experimental study. Sci Rep. 2023;13(1):9423.

22. Altmann CH, Zvonova E, Richter L, Schuller PO. Pulmonary recovery directly after COVID-19 and in Long-COVID. Respir Physiol Neurobiol. 2023;315:104112.

23. Garcia-Molina A, Garcia-Carmona S, Espina-Bou M, Rodriguez-Rajo P, Sanchez-Carrion R, Ensenat-Cantallops A. [Neuropsychological rehabilitation for post-COVID-19 syndrome: Results of a clinical program and six-month follow up.]. Neurologia. 2022.

24. Derksen C, Rinn R, Gao L, Dahmen A, Cordes C, Kolb C, et al. Longitudinal Evaluation of an Integrated Post-COVID-19/Long COVID Management Program Consisting of Digital Interventions and Personal Support: Randomized Controlled Trial. J Med Internet Res. 2023;25:e49342.

25. Sathyamoorthy M, Verduzco-Gutierrez M, Varanasi S, Ward R, Spertus J, Shah S. Enhanced external counterpulsation for management of symptoms associated with long COVID. Am Heart J Plus. 2022;13:100105.

26. Ghali A, Lacombe V, Ravaiau C, Delattre E, Ghali M, Urbanski G, et al. The relevance of pacing strategies in managing symptoms of post-COVID-19 syndrome. J Transl Med. 2023;21(1):375.

27. Tanguay P, Gaboury I, Daigle F, Bhéreur A, Dubois O, Lagueux E, et al. Post-exertional malaise may persist in Long COVID despite learning STOP-REST-PACE. Fatigue: Biomedicine, Health & Behavior. 2023;11:1–16.

28. Hasenoehrl T, Palma S, Huber DF, Kastl S, Steiner M, Jordakieva G, et al. Post-COVID: effects of physical exercise on functional status and work ability in health care personnel. Disabil Rehabil. 2023;45(18):2872–8.

29. Oka T. A patient who recovered from post-COVID myalgic encephalomyelitis/chronic fatigue syndrome: a case report. Biopsychosoc Med. 2023;17(1):8.

30. (NICE) NIfHaCE. COVID-19 rapid guideline: managing the long-term effects of COVID-19. National Institute for Health and Care Excellence (NICE); 2020 December 18 2020.

31. Muller K, Poppele I, Ottiger M, Zwingmann K, Berger I, Thomas A, et al. Impact of Rehabilitation on Physical and Neuropsychological Health of Patients Who Acquired COVID-19 in the Workplace. Int J Environ Res Public Health. 2023;20(2).

32. Wong J, Kudla, A., Ezeife, N., Crown, D., Capraro, P., Trierweiler, R., Tomazin, S., Heinemann, A. W., & Pham, T. Lessons Learned by Rehabilitation Counselors and Physicians in Services to COVID-19 Long-Haulers: A Qualitative Study. Rehabilitation Counseling Bulletin. 2022;66(1):25–35.

33. Returning safely to work after long COVID. British Journal of Healthcare Assistants. 2022;16(1):50-.

34. Burton K, Caine, A., Macniven, L., Porter, S., Rayner, C., & Yarker, J. COVID-19 return to work guide: for managers. Society of Occupational Medicine. 2021.

35. Medicine TSoO. SOM issues long Covid and Covid return-to-work guidance. 2021:8.

36. Madan I, Briggs T, Chew-Graham C. Supporting patients with long COVID return to work. Br J Gen Pract. 2021;71(712):508–9.

37. Baril R, Clarke J, Friesen M, Stock S, Cole D. Management of return-to-work programs for workers with musculoskeletal disorders: a qualitative study in three Canadian provinces. Social Science & Medicine. 2003;57(11):2101–14.

38. Lunt J, Hemming S, Elander J, Baraniak A, Burton K, Ellington D. Experiences of workers with post-COVID-19 symptoms can signpost suitable workplace accommodations. International Journal of Workplace Health Management. 2022;15(3):359–74.

39. Macdonald EL, Drushca; Raynor, Clare; Yarker, Jo;. COVID-19 infection and long COVID – guide for managers. European Agency for Health and Safety at Work (EUOSHA) 2021. 2021.

40. Nurek M, Rayner C, Freyer A, Taylor S, Järte L, MacDermott N, et al. Recommendations for the recognition, diagnosis, and management of long COVID: a Delphi study. Br J Gen Pract. 2021;71(712):e815–e25.

41. Rajan S, Khunti K, Alwan N, Steves C, MacDermott N, Morsella A, et al. European Observatory Policy Briefs. In the wake of the pandemic: Preparing for Long COVID. Copenhagen (Denmark): European Observatory on Health Systems and Policies © World Health Organization 2021 (acting as the host organization for, and secretariat of, the European Observatory on Health Systems and Policies). 2021.

42. Shah W, Hillman T, Playford ED, Hishmeh L. Managing the long term effects of covid-19: summary of NICE, SIGN, and RCGP rapid guideline. Bmj. 2021;372:n136.

43. Stewart-Patterson C, Bourgeois R, Martin DW. The Importance of Keeping Patients with Post-Acute Sequelae of SARS-CoV-2 Infection (Long COVID) Engaged in Work. Am Fam Physician. 2021;103(12):710.

44. Beauchamp MK, Janaudis-Ferreira T, Wald J, Aceron R, Bhutani M, Bourbeau J, et al. Canadian Thoracic Society position statement on rehabilitation for COVID-19 and implications for pulmonary rehabilitation. Canadian Journal of Respiratory, Critical Care, and Sleep Medicine. 2022;6(1):9–13.

45. Moulaei K, Sheikhtaheri A, Fatehi F, Shanbehzadeh M, Bahaadinbeigy K. Patients’ perspectives and preferences toward telemedicine versus in-person visits: a mixed-methods study on 1226 patients. BMC Med Inform Decis Mak. 2023;23(1):261.

