## Supplemental Appendix 1 for "Return-to-work for People Living with Long COVID: A Scoping Review of Interventions and Recommendations"

**Appendix A – Full Search Strategy**

**Ovid MEDLINE(R) ALL <1946 to October 05, 2023>**

Date searched: Oct 6, 2023

Results: 473

1	Post-Acute COVID-19 Syndrome/	2482

2	((long or (long term not long term care) or long haul* or longterm or longhaul or post acute or postacute or after acute or sequela* or protracted or post-infect* or post-viral or post-discharg* or non-recover* or nonrecover* or PASC or chronic or persist* or linger* or continuing or continual) adj5 (COVID or COVID-19 or COVID19 or coronavirus* or corona virus* or 2019-nCoV or 19nCoV or 2019nCoV or nCoV or n-CoV or SARS-CoV-2 or SARS-CoV2 or SARSCoV-2 or SARSCoV2 or 2019-novel CoV or Sars-coronavirus2 or novel CoV)).mp.	15806

3	((long or long term or long haul* or longterm or longhaul or ongoing or chronic or lengthy or protracted or persist* or linger* or continuing or continual* or post-acute or postacute or post-viral or post-discharg* or post-infect* or residual) adj2 (outcomes or symptom* or morbidity or manifestation* or issues or effects or difficulties or challeng* or problems or complications or disturbances or consequences or impairments or dysfunction or function or functioning or functional or abnormalities or dizziness or headache* or dyspnea or fatigue or breath or breathing or lung or respiratory or tachycardia or palpitation* or neuro* or concentration or concentrating or brain fog or cough or ache* or pain* or taste or smell or olfactory or olfaction or gustatory)).mp. and (COVID or COVID-19 or COVID19 or coronavirus* or corona virus* or 2019-nCoV or 19nCoV or 2019nCoV or nCoV or n-CoV or SARS-CoV-2 or SARS-CoV2 or SARSCoV-2 or SARSCoV2 or 2019-novel CoV or Sars-coronavirus2 or novel CoV).tw.	10068

4	((postcovid or postcorona* or post-COVID or post-COVID-19 or post-COVID19 or post-coronavirus* or post-corona virus*) adj3 (syndrome or outcomes or symptom* or morbidity or manifestation* or issues or effects or difficulties or challeng* or problems or complications or disturbances or consequences or impairments or dysfunction or function or functioning or functional or abnormalities or dizziness or headache* or dyspnea or fatigue or breath or breathing or lung or respiratory or tachycardia or palpitation* or neuro* or concentration or concentrating or brain fog or cough or ache* or pain* or taste or smell or olfactory or olfaction or gustatory)).mp.	2486

5	1 or 2 or 3 or 4	21989

6	((work* or job or jobs or employee*) adj3 (modification* or modified or accommodat*)).mp.	4881

7	("occupational rehab*" or "vocational rehab*" or "workplace intervention*").mp.	4592

8	((work or worker* or workplace or employ* or occupation or vocational) and (rehab* or telerehab* or mindfulness or "acceptance and commitment therap*" or "relaxation technique*" or "relaxation therap*" or "stop rest pace" or "breathing technique*")).mp.	55285

9	Post-Acute COVID-19 Syndrome/rh, th [Rehabilitation, Therapy]	16

10	(Return* to work* or "sick* leave*" or "worker* compensation" or WCB or "disability coverage").mp.	32779

11	((work* or job or jobs or employee* or personnel or professionals) adj3 (perform* or "carry* out" or abilit* or inabilit* or able or return* or reintegrat*)).mp.	73295

12	((work or worker* or workplace or employ* or job or jobs or personnel or professionals or occupation* or vocational) and ("push-crash cycle" or "boom-bust cycle" or "autonomic dysfunction*" or "post exertional malaise" or PEM or "post exertional symptom exacerbation*" or PESE or "short term disability" or "longterm disability" or "long term disability" or absenteeism or presenteeism)).mp.	12496

13	((work* or employee* or job* or occupational or vocational) adj3 (performance or limitation* or functioning or efficiency or efficacy or productivity or capacity)).mp.	60885

14	6 or 7 or 8 or 9 or 10 or 11 or 12 or 13	180532

15	5 and 14	473

**Embase <1974 to 2023 October 05> (OVID Interface)**

Date searched: Oct 6, 2023

Results: 1028

1	long COVID/	5679

2	((long or (long term not long term care) or long haul* or longterm or longhaul or post acute or postacute or after acute or sequela* or protracted or post-infect* or post-viral or post-discharg* or non-recover* or nonrecover* or PASC or chronic or persist* or linger* or continuing or continual) adj5 (COVID or COVID-19 or COVID19 or coronavirus* or corona virus* or 2019-nCoV or 19nCoV or 2019nCoV or nCoV or n-CoV or SARS-CoV-2 or SARS-CoV2 or SARSCoV-2 or SARSCoV2 or 2019-novel CoV or Sars-coronavirus2 or novel CoV)).mp.	20498

3	((long or long term or long haul* or longterm or longhaul or ongoing or chronic or lengthy or protracted or persist* or linger* or continuing or continual* or post-acute or postacute or post-viral or post-discharg* or post-infect* or residual) adj2 (outcomes or symptom* or morbidity or manifestation* or issues or effects or difficulties or challeng* or problems or complications or disturbances or consequences or impairments or dysfunction or function or functioning or functional or abnormalities or dizziness or headache* or dyspnea or fatigue or breath or breathing or lung or respiratory or tachycardia or palpitation* or neuro* or concentration or concentrating or brain fog or cough or ache* or pain* or taste or smell or olfactory or olfaction or gustatory)).mp. and (COVID or COVID-19 or COVID19 or coronavirus* or corona virus* or 2019-nCoV or 19nCoV or 2019nCoV or nCoV or n-CoV or SARS-CoV-2 or SARS-CoV2 or SARSCoV-2 or SARSCoV2 or 2019-novel CoV or Sars-coronavirus2 or novel CoV).tw.	21005

4	((postcovid or postcorona* or post-COVID or post-COVID-19 or post-COVID19 or post-acute-covid* or post-coronavirus* or post-corona virus*) adj3 (syndrome or outcomes or symptom* or morbidity or manifestation* or issues or effects or difficulties or challeng* or problems or complications or disturbances or consequences or impairments or dysfunction or function or functioning or functional or abnormalities or dizziness or headache* or dyspnea or fatigue or breath or breathing or lung or respiratory or tachycardia or palpitation* or neuro* or concentration or concentrating or brain fog or cough or ache* or pain* or taste or smell or olfactory or olfaction or gustatory)).mp.	3953

5	1 or 2 or 3 or 4	35523

6	Return to Work/	9744

7	work capacity/	13821

8	job accommodation/ or work resumption/	3954

9	vocational rehabilitation/	8825

10	((work* or job or jobs or employee*) adj3 (modification* or modified or accommodat*)).mp.	6384

11	("occupational rehab*" or "vocational rehab*" or "workplace intervention*").mp.	11520

12	((work or worker* or workplace or employ* or occupation or vocational) and (rehab* or telerehab* or mindfulness or "acceptance and commitment therap*" or "relaxation technique*" or "relaxation therap*" or "stop rest pace" or "breathing technique*")).mp.	69992

13	exp long COVID/rh, th [Rehabilitation, Therapy]	198

14	(Return* to work* or "sick* leave*" or "worker* compensation" or WCB or "disability coverage").mp.	33590

15	((work* or job or jobs or employee* or personnel or professionals) adj3 (perform* or "carry* out" or abilit* or inabilit* or able or return* or reintegrat*)).mp.	104769

16	((work or worker* or workplace or employ* or job or jobs or personnel or professionals or occupation* or vocational) and ("push-crash cycle" or "boom-bust cycle" or "autonomic dysfunction*" or "post exertional malaise" or PEM or "post exertional symptom exacerbation*" or PESE or "short term disability" or "longterm disability" or "long term disability" or absenteeism or presenteeism)).mp.	21671

17	((work* or employee* or job* or occupational or vocational) adj3 (performance or limitation* or functioning or efficiency or efficacy or productivity or capacity)).mp.	89232

18	or/6-17	240022

19	5 and 18	1028

**APA PsycInfo <1806 to October Week 1 2023> (Ovid Interface)**

Date searched: Oct 6, 2023

Results: 57

1	post-covid-19 conditions/	37

2	((long or (long term not long term care) or long haul* or longterm or longhaul or post acute or postacute or after acute or sequela* or protracted or post-infect* or post-viral or post-discharg* or non-recover* or nonrecover* or PASC or chronic or persist* or linger* or continuing or continual) adj5 (COVID or COVID-19 or COVID19 or coronavirus* or corona virus* or 2019-nCoV or 19nCoV or 2019nCoV or nCoV or n-CoV or SARS-CoV-2 or SARS-CoV2 or SARSCoV-2 or SARSCoV2 or 2019-novel CoV or Sars-coronavirus2 or novel CoV)).mp.	1164

3	((long or long term or long haul* or longterm or longhaul or ongoing or chronic or lengthy or protracted or persist* or linger* or continuing or continual* or post-acute or postacute or post-viral or post-discharg* or post-infect* or residual) adj2 (outcomes or symptom* or morbidity or manifestation* or issues or effects or difficulties or challeng* or problems or complications or disturbances or consequences or impairments or dysfunction or function or functioning or functional or abnormalities or dizziness or headache* or dyspnea or fatigue or breath or breathing or lung or respiratory or tachycardia or palpitation* or neuro* or concentration or concentrating or brain fog or cough or ache* or pain* or taste or smell or olfactory or olfaction or gustatory)).mp. and (COVID or COVID-19 or COVID19 or coronavirus* or corona virus* or 2019-nCoV or 19nCoV or 2019nCoV or nCoV or n-CoV or SARS-CoV-2 or SARS-CoV2 or SARSCoV-2 or SARSCoV2 or 2019-novel CoV or Sars-coronavirus2 or novel CoV).tw.	1020

4	((postcovid or postcorona* or post-COVID or post-COVID-19 or post-COVID19 or post-coronavirus* or post-corona virus*) adj3 (syndrome or outcomes or symptom* or morbidity or manifestation* or issues or effects or difficulties or challeng* or problems or complications or disturbances or consequences or impairments or dysfunction or function or functioning or functional or abnormalities or dizziness or headache* or dyspnea or fatigue or breath or breathing or lung or respiratory or tachycardia or palpitation* or neuro* or concentration or concentrating or brain fog or cough or ache* or pain* or taste or smell or olfactory or olfaction or gustatory)).mp.	158

5	1 or 2 or 3 or 4	1844

6	reemployment/	1906

7	job performance/ or employee productivity/	23578

8	employee absenteeism/	2437

9	vocational rehabilitation/	6398

10	((work* or job or jobs or employee*) adj3 (modification* or modified or accommodat*)).mp.	2119

11	("occupational rehab*" or "vocational rehab*" or "workplace intervention*").mp.	9860

12	((work or worker* or workplace or employ* or occupation or vocational) and (rehab* or telerehab* or mindfulness or "acceptance and commitment therap*" or "relaxation technique*" or "relaxation therap*" or "stop rest pace" or "breathing technique*")).mp.	31183

13	(Return* to work* or "sick* leave*" or "worker* compensation" or WCB or "disability coverage").mp.	7269

14	((work* or job or jobs or employee* or personnel or professionals) adj3 (perform* or "carry* out" or abilit* or inabilit* or able or return* or reintegrat*)).mp.	55030

15	((work or worker* or workplace or employ* or job or jobs or personnel or professionals or occupation* or vocational) and ("push-crash cycle" or "boom-bust cycle" or "autonomic dysfunction*" or "post exertional malaise" or PEM or "post exertional symptom exacerbation*" or PESE or "short term disability" or "longterm disability" or "long term disability" or absenteeism or presenteeism)).mp.	5834

16	((work* or employee* or job* or occupational or vocational) adj3 (performance or limitation* or functioning or efficiency or efficacy or productivity or capacity)).mp.	59784

17	((work* or employee* or job* or occupational or vocational) adj3 (performance or limitation* or functioning or efficiency or efficacy or productivity or capacity)).mp.	59784

18	or/6-17	114519

19	5 and 18	57

**CINAHL Plus with Full Text (EBSCOhost Interface)**

Date searched: Oct 6, 2023

Results: 204

S1: (MH "Post-Acute COVID-19 Syndrome") OR ( ((long or (long-term not long-term-care) or long-haul* or longterm or longhaul or post-acute or postacute or after-acute or sequela* or protracted or post-infect* or post-viral or post-discharg* or non-recover* or nonrecover* or PASC or chronic or persist* or linger* or continuing or continual) N5 (COVID or COVID-19 or COVID19 or coronavirus* or corona-virus* or 2019-nCoV or 19nCoV or 2019nCoV or nCoV or n-CoV or SARS-CoV-2 or SARS-CoV2 or SARSCoV-2 or SARSCoV2 or 2019-novel-CoV or Sars-coronavirus2 or novel CoV)) ) OR ( ((long or long-term or long-haul* or longterm or longhaul or ongoing or chronic or lengthy or protracted or persist* or linger* or continuing or continual* or post-acute or postacute or post-viral or post-discharg* or post-infect* or residual) N2 (outcomes or symptom* or morbidity or manifestation* or issues or effects or difficulties or challeng* or problems or complications or disturbances or consequences or impairments or dysfunction or function or functioning or functional or abnormalities or dizziness or headache* or dyspnea or fatigue or breath or breathing or lung or respiratory or tachycardia or palpitation* or neuro* or concentration or concentrating or brain-fog or cough or ache* or pain* or taste or smell or olfactory or olfaction or gustatory)) AND (COVID or COVID-19 or COVID19 or coronavirus* or corona-virus* or 2019-nCoV or 19nCoV or 2019nCoV or nCoV or n-CoV or SARS-CoV-2 or SARS-CoV2 or SARSCoV-2 or SARSCoV2 or 2019-novel-CoV or Sars-coronavirus2 or novel CoV) ) OR ( ((postcovid or postcorona* or post-COVID or post-COVID-19 or post-COVID19 or post-coronavirus* or post-corona-virus* or post-acute-covid*) N3 (syndrome or outcomes or symptom* or morbidity or manifestation* or issues or effects or difficulties or challeng* or problems or complications or disturbances or consequences or impairments or dysfunction or function or functioning or functional or abnormalities or dizziness or headache* or dyspnea or fatigue or breath or breathing or lung or respiratory or tachycardia or palpitation* or neuro* or concentration or concentrating or brain-fog or cough or ache* or pain* or taste or smell or olfactory or olfaction or gustatory)) )

S2: (MH "Job Accommodation") or (MH "Job Re-Entry") OR (MH "Job Performance")

S3: ((work* or job or jobs or employee*) N3 (modification* or modified or accommodat*)) ) OR "occupational rehab*" or "vocational rehab*" or "workplace intervention*" or "Return* to work*" or "sick* leave*" or "worker* compensation" or WCB or "disability coverage"

S4	((work or worker* or workplace or employ* or occupation or vocational) and (rehab* or telerehab* or mindfulness or "acceptance and commitment therap*" or "relaxation technique*" or "relaxation therap*" or "stop rest pace" or "breathing technique*"))

S5	((work* or job or jobs or employee* or personnel or professionals) N3 (perform* or "carry* out" or abilit* or inabilit* or able or return* or reintegrat*))

S6	((work or worker* or workplace or employ* or job or jobs or personnel or professionals or occupation* or vocational) and ("push-crash cycle" or "boom-bust cycle" or "autonomic dysfunction*" or "post exertional malaise" or PEM or "post exertional symptom exacerbation*" or PESE or "short term disability" or "longterm disability" or "long term disability" or absenteeism or presenteeism))

S7	((work* or employee* or job* or occupational or vocational) N3 (performance or limitation* or functioning or efficiency or efficacy or productivity or capacity))

S8	S1 AND (S2 OR S3 OR S4 OR S5 OR S6 OR S7)

**Cochrane Library** - Trials database only (Wiley Interface)

Date searched: Oct 6, 2023

Results: ( Trials =90 )

#1	[mh ^"Post-Acute COVID-19 Syndrome"]

#2	((long or long-term or long-haul or long-hauler or longterm or longhaul or post-acute or postacute or after-acute or sequela* or protracted or post-infection or post-viral or post-discharge or non-recovered or nonrecover* or PASC or chronic or persist* or linger* or continuing or continual) NEAR/5 (COVID or COVID-19 or COVID19 or coronavirus* or corona-virus or "2019-nCoV" or "19nCoV" or "2019nCoV" or nCoV or n-CoV or SARS-CoV-2 or SARS-CoV2 or SARSCoV-2 or SARSCoV2 or "2019-novel-CoV" or Sars-coronavirus2 or novel-CoV)):ti,ab,kw

#3	((long or long-term or long-haul or long-hauler or longterm or longhaul or ongoing or chronic or lengthy or protracted or persist* or linger* or continuing or continual* or post-acute or postacute or post-viral or post-discharge or post-infection or residual) NEAR/2 (outcomes or symptom* or morbidity or manifestation* or issues or effects or difficulties or challeng* or problems or complications or disturbances or consequences or impairments or dysfunction or function or functioning or functional or abnormalities or dizziness or headache* or dyspnea or fatigue or breath or breathing or lung or respiratory or tachycardia or palpitation* or neuro* or concentration or concentrating or brain-fog or cough or ache* or pain* or taste or smell or olfactory or olfaction or gustatory)):ti,ab,kw and (COVID or COVID-19 or COVID19 or coronavirus* or corona-virus or "2019-nCoV" or "19nCoV" or "2019nCoV" or nCoV or n-CoV or SARS-CoV-2 or SARS-CoV2 or SARSCoV-2 or SARSCoV2 or "2019-novel-CoV" or Sars-coronavirus2 or novel-CoV):ti,ab,kw

#4	((postcovid or postcorona* or post-COVID or post-COVID-19 or post-COVID19 or post-coronavirus or post-corona-virus or post-acute-covid) NEAR/3 (syndrome or outcomes or symptom* or morbidity or manifestation* or issues or effects or difficulties or challeng* or problems or complications or disturbances or consequences or impairments or dysfunction or function or functioning or functional or abnormalities or dizziness or headache* or dyspnea or fatigue or breath or breathing or lung or respiratory or tachycardia or palpitation* or neuro* or concentration or concentrating or brain-fog or cough or ache* or pain* or taste or smell or olfactory or olfaction or gustatory)):ti,ab,kw

#5	(#1 OR #2 OR #3 OR #4)

#6	((work* or job or jobs or employee*) NEAR/3 (modification* or modified or accommodat*)):ti,ab,kw

#7	("occupational rehabilitation" or "vocational rehabilitation" or "workplace intervention"):ti,ab,kw

#8	((work or worker* or workplace or employ* or occupation or vocational) and (rehab* or telerehab* or mindfulness or "acceptance and commitment therapy" or "relaxation technique" or "relaxation therapy" or "stop rest pace" or "breathing technique")):ti,ab,kw

#9	[mh ^"Post-Acute COVID-19 Syndrome"/rh,th]

#10	(Return-to-work or "sick leave" or (worker* NEXT compensation) or WCB or "disability coverage"):ti,ab,kw

#11	((work* or job or jobs or employee* or personnel or professionals) NEAR/3 (perform* or "carry out" or "carrying out" or abilit* or inabilit* or able or return* or reintegrat*)):ti,ab,kw

#12	((work or worker* or workplace or employ* or job or jobs or personnel or professionals or occupation* or vocational) and ("push-crash cycle" or "boom-bust cycle" or "autonomic dysfunction" or "post exertional malaise" or PEM or "post exertional symptom exacerbation" or PESE or "short term disability" or "longterm disability" or "long term disability" or absenteeism or presenteeism)):ti,ab,kw

#13	((work* or employee* or job* or occupational or vocational) NEAR/3 (performance or limitation* or functioning or efficiency or efficacy or productivity or capacity)):ti,ab,kw

#14	#6 OR #7 OR #8 OR #9 OR #10 OR #11 OR #12 OR #13

#15	#5 AND #14

Grey literature search

**Government of Canada Publications** (<https://www.publications.gc.ca/site/eng/home.html>) - Searched Nov 7 (post-covid work; long-covid work) (0 useful results)

**Custom Search Engine for Canadian Public Health Information** (<https://www.ophla.ca/p/customsearchcanada.html>)

Searched November 7, 2023

"return to work" long-covid OR post-covid-syndrome OR post-covid-condition OR post-covid-19-condition OR post-covid-19-syndrome (retrieved 24 results)

**Custom Search Engine for US State Government Information (**[**https://www.ophla.ca/p/customsearchusstates.html**](https://www.ophla.ca/p/customsearchusstates.html)**)**

Searched November 7, 2023

"return to work" long-covid OR post-covid-syndrome OR post-covid-condition OR post-covid-19-condition OR post-covid-19-syndrome (only retrieved 11 results)

**MedRxiv (**[**https://www.medrxiv.org/**](https://www.medrxiv.org/)**) (USE ADVANCED SEARCH,** Searched in title/abstract**)**

Searched November 7, 2023

(total 7 results)

Long-covid return-to-work (all terms) - 5 results

post-covid return-to-work (all terms) - 3 results

**Bielefeld Academic Search Engine (BASE) (https://www.base-search.net/)**

Searched November 7, 2023

Searched using "entire document" and "verbatim search"

"Return to work" long-covid - 75 results

"Return to work" post-covid 110 results

**OAIster (oaister.on.worldcat.org) - Use Advanced search**

Searched November 7, 2023 (Total 12 items downloaded)

"Return to work" long-covid

"Return to work" post-covid

**Google (Advanced search) (searched with an incognito browser):**

Searched Nov 7, 2023

"return to work" long-covid OR post-covid-syndrome OR post-covid-condition OR post-covid-19-condition OR post-covid-19-syndrome filetype:pdf (review 168 results)

"return to work" long-covid OR post-covid-syndrome OR post-covid-condition OR post-covid-19-condition OR post-covid-19-syndrome -filetype:pdf -site:pubmed.ncbi.nlm.nih.gov -Oxford -elsevier -science-direct -sage -wiley (review first 100 results)

"Workers Compensation" long-covid OR post-covid-syndrome OR post-covid-condition OR post-covid-19-condition OR post-covid-19-syndrome (review first 50 results)

**These websites were scanned for any relevant documents on Nov 7, 2023:**

**Alberta Health Services "Recovery & Rehabilitation After COVID-19: Resources for Health Professionals**

[**https://www.albertahealthservices.ca/topics/Page17540.aspx**](https://www.albertahealthservices.ca/topics/Page17540.aspx)

**WHO websites relevant to long covid**

Rehabilitation and Covid-19 <https://www.who.int/teams/noncommunicable-diseases/covid-19/rehabilitation>

Post covid condition

<https://www.who.int/teams/health-care-readiness/post-covid-19-condition>

High-level meeting on post-COVID conditions (‎long COVID)‎: a virtual meeting hosted by the WHO Regional Office for Europe, 19 March 2021

<https://www.who.int/europe/publications/i/item/WHO-EURO-2021-2410-42165-58100>
